# Multiday rhythms shape mood dynamics in depression

**DOI:** 10.64898/2026.08.25.26361076

**Authors:** Nandini P. Sekar, Joline M. Fan, Kristin K. Sellers, Daniela Astudillo Maya, Alexandra Tremblay-McGaw, Natalie Becker, Alice Le Berre, Anusha Allawala, Elissa Hamlat, Leo P. Sugrue, Vikram R. Rao, Andrew D. Krystal, Edward F. Chang, Ankit N. Khambhati

## Abstract

Mood fluctuations in major depressive disorder are difficult to anticipate. The biological neural rhythms that organize mood dynamics over days to weeks remain unknown. In individuals implanted with a chronic neural sensing and stimulation device for treatment-resistant depression, we collected years-long intracranial neural recordings alongside daily mood ratings. Both mood and limbic neural activity fluctuated cyclically with multiday (multidien) periodicities of 2–34 days. An individual’s daily phase position within mood cycles tracked depression severity, distinguishing whether symptoms were rising, peaking, or resolving. Neural rhythms led mood cycles and forecast an individual’s mood trajectory up to 30 days in advance, outperforming models based on raw neural activity. Electrical stimulation reshaped these rhythms, shifting individuals away from the peak-depression phase of their multidien cycle. Our results identify multidien rhythms as an organizing principle of mood in depression and a forecastable, modifiable target for chronotherapeutic neuromodulation.

## Introduction

Mood is a dynamic internal state driven by underlying brain activity. It rises and falls over hours, days, and weeks, and profoundly shapes how a person thinks, experiences, and interacts with the world. In health, fluctuations in internal state are often adaptive, allowing internal state to track changing biological and environmental demands^1^. In depression, however, mood dynamics can become pathological, where low mood may persist, worsen, or recur in ways that are difficult to anticipate^2,3^. Fluctuations in internal state may create periods where mood symptoms are heightened. Understanding the temporal architecture of mood is therefore central to understanding depression itself. Yet, despite decades of work on depressive symptoms and episodes, little is known about the biological rhythms that organize mood over time, or the neural dynamics from which they emerge.

Multiday rhythms are increasingly recognized as a widespread biological principle, with slow cyclical organisation of the brain, body, and behavior being observed across populations. Multiday rhythms appear in heart rate^4^ and smartphone usage^5^ in non-clinical populations. In epilepsy, such rhythms modulate seizure likelihood^6–9^. Rhythmic cycles spanning days have also been reported in mood in bipolar disorder and depression^10,11^, and slow changes in neural dynamics have been associated with depression recovery^12,13^, suggesting that mood symptoms and the underlying neural activity shaping them may fluctuate across slow, multiday timescales. We therefore hypothesized that depression is organized, in part, by multiday neural rhythms that shape mood dynamics across recurring windows of vulnerability and resilience.

We tested this hypothesis using chronic intracranial electroencephalography (iEEG) recordings from people with treatment-resistant depression (TRD) enrolled in the PRESIDIO clinical trial (NCT04004169) and implanted with the NeuroPace Responsive Neurostimulation (RNS) System. The RNS System enabled years-long recordings from limbic brain regions alongside daily self-reported mood in natural, ambulatory settings (Fig.1a). Using chronic iEEG signals and mood ratings, we identified multiday cycles in mood and corresponding multiday oscillatory structure in the neural local field potential (Fig.1b). These chronic recordings revealed an extension of the traditional power spectrum into ultra-slow frequencies below the delta band (Fig.1c), reaching timescales of days to months (∼10^−6^ Hz; Fig.1d). We show that these ultra-slow iEEG rhythms drive cyclical fluctuations in mood symptoms through rising, falling, peak, and low phases–enabling forecasting of mood trajectory days in advance (Fig.1e). Finally, using the stimulation capabilities of the RNS System, we find that neural multiday rhythms are modifiable and that their timescales change in response to electrical stimulation (Fig.1f). Together, these findings support rhythm-informed strategies for anticipating elevated mood symptoms and responsively suppressing them before they occur.

**Figure 1:**
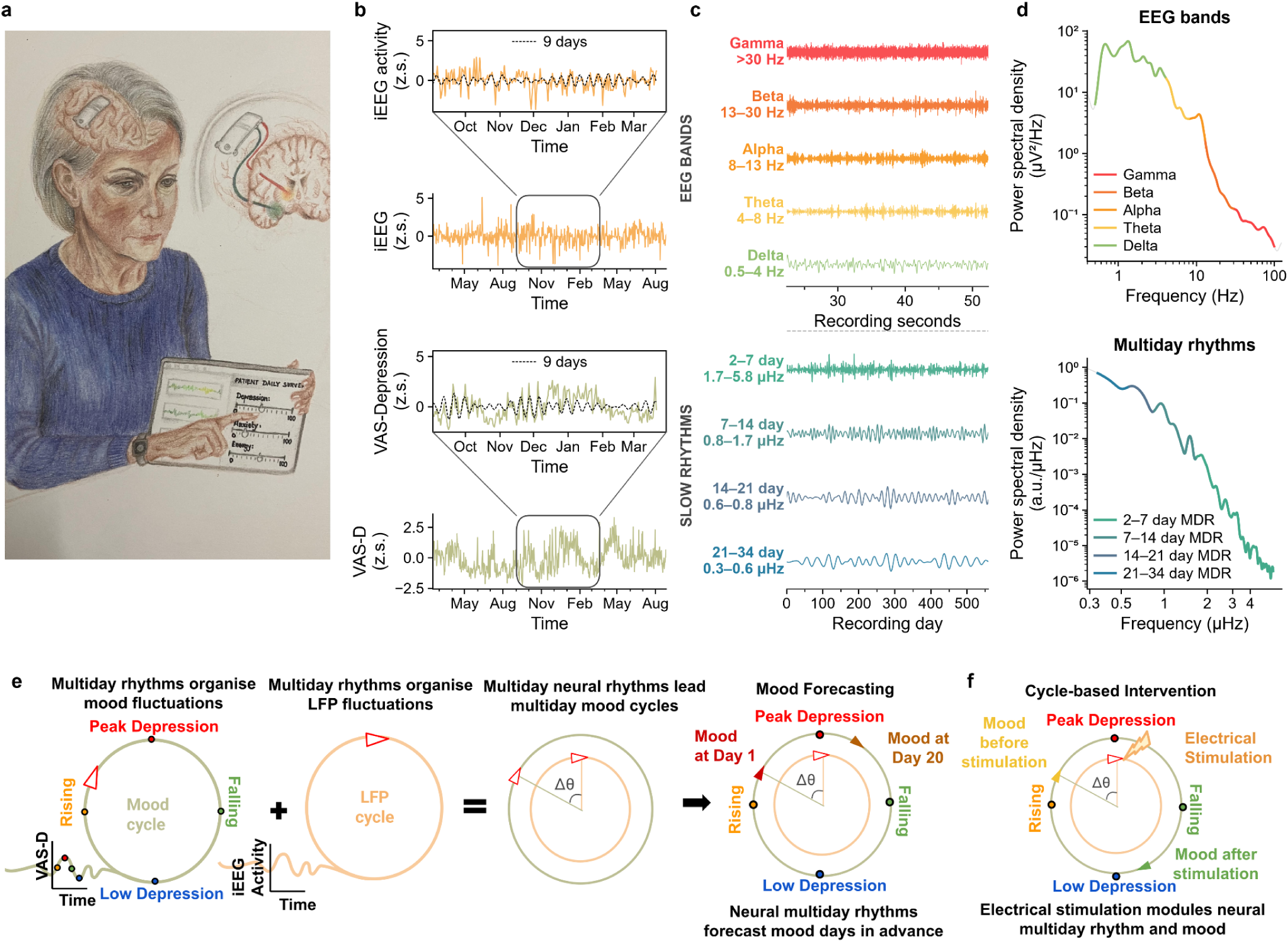
Multiday rhythms organize fluctuations in iEEG and behavior. **a.** Representative research participant implanted with the NeuroPace RNS System and providing chronic intracranial EEG (iEEG) recordings and daily mood symptom ratings. **b.** Representative time-series of iEEG activity (top, orange) and self-reported mood (VAS-Depression, bottom, green) from one participant. Inset (right): zoomed window showing each signal overlaid with its 9-day wavelet reconstruction (dashed, black; iEEG: r = 0.40, p < 0.001; VAS-Depression: r = 0.41, p < 0.001). Scale bars: 60 days (horizontal), 0.25 z.s. (vertical). **c**. iEEG signal filtered in canonical frequency bands over a 30-second snapshot (top) and in multiday frequency bands over 560 days (bottom), showing time-varying rhythmic structure, in a single participant. **d.** Power spectral density of the iEEG signal in panel c, showing traditional EEG frequency bands (top) and multiday slow rhythm bands (2-7, 7-14, 14-21, 21-34 days; bottom). Multiday bands shown for visualization only; analyses use the full continuous wavelet spectrum (2-34 days). **e.** Hypothesized coordination of multiday mood rhythm by the local field potential rhythm. Trajectory of the multiday rhythms comprises four phases: rising, peak, falling, and low mood. Cycling in the local field potential leads to cycling of mood–enabling the forecasting of mood trajectory days in advance. **f.** Hypothesized cycle-based intervention showing electrical stimulation modulating multiday rhythm of the local field potential and thereby regulating fluctuation in mood. Panels b-d show representative data from one participant and one brain region in the limbic network.

## Results

Five participants with TRD (Extended Data Table 1) were implanted with the RNS System, each with intracranial leads sampling two of the following regions: orbitofrontal cortex, amygdala, subgenual cingulate, hypothalamus, thalamus, hippocampus, nucleus accumbens, and anterior limb of the internal capsule (Extended Data Fig.1). Each four-contact lead was sampled as two bipolar-referenced channels, yielding four channels of iEEG per participant. Lead implant locations were optimized per participant using a personalized, in-patient brain mapping approach^14,15^ that yielded brain regions that demonstrated the best therapeutic stimulation response and the most informative acute biomarker recordings (Extended Data Table 2). iEEG and self-reported Visual Analogue Scale of Depression (VAS-D) were sampled multiple times a day over years (1.47 ± 0.50 years, range: 0.67–1.95 years). Clinical depression severity was evaluated through Montgomery-Asberg Depression Rating Scale (MADRS) administered roughly every two weeks (14.5 ± 4.7 days, range: 9.5–19.8 days). In this study, we operationalized mood as the self-reported VAS-D (0–100), and clinical depression severity as the clinician-administered MADRS. VAS-D provides dense temporal resolution of moment-to-moment mood dynamics, while MADRS provides a periodic (∼biweekly) clinical anchor that summarizes the symptom state.

**Extended Data Figure 1:**
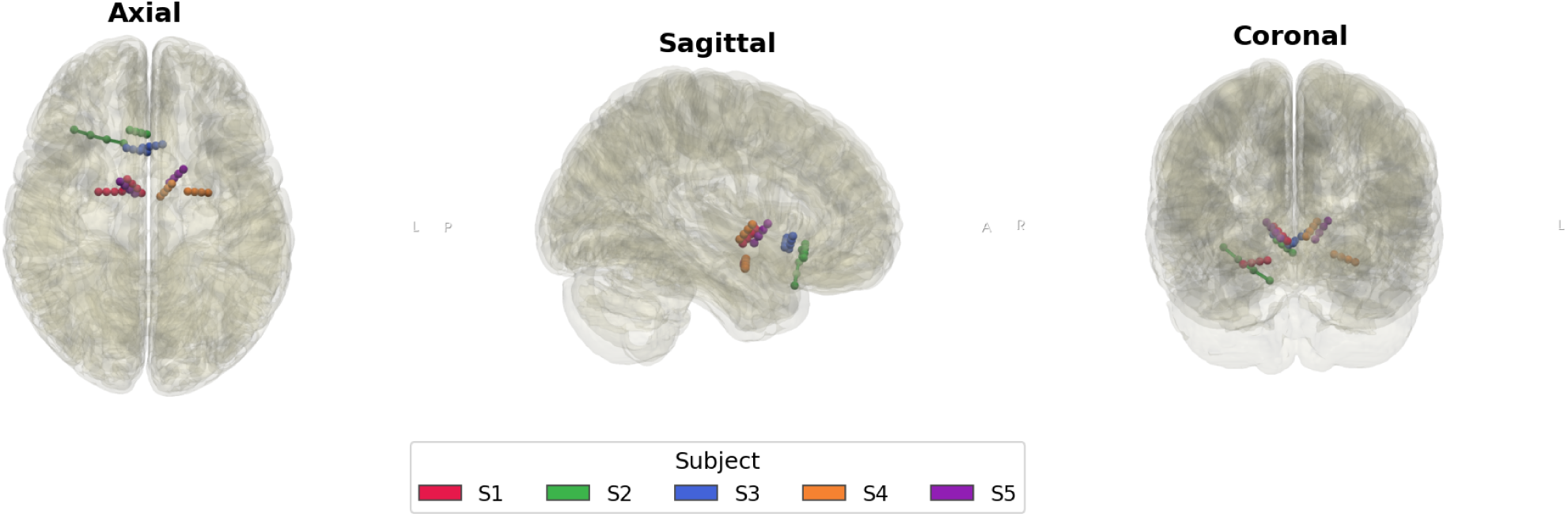
Electrode localization per participant.

**Extended Data Table 1:** Demographic and clinical profiles of study participants.

| Participant | Primary & comorbid diagnoses | Suicidality history | Predominant presenting symptoms |
| --- | --- | --- | --- |
| S1 (41-50, F) | MDD; dyslexia; GAD | History of suicide attempt | Melancholy, hopelessness, insomnia |
| S2 (31-40, M) | MDD (melancholic); insomnia; anxiety symptoms | Chronic SI | Fatigue, psychomotor symptoms, hopelessness |
| S3 (31-40, F) | MDD; GAD; SAD; panic symptoms | History of suicide attempt | Loss of appetite, anhedonia, dread |
| S4 (31-40, F) | MDD; GAD; insomnia; PMDD traits; ND traits | Chronic SI | Apathy, insomnia, hopelessness |
| S5 (41-50, M) | MDD; SAD; PTSD symptoms | Passive SI | Anhedonia, guilt, poor energy |

Abbreviations: F, female; GAD, generalized anxiety disorder; M, male; MDD, major depressive disorder; MDE, major depressive episode; ND, neurodevelopmental; PMDD, premenstrual dysphoric disorder; PTSD, post-traumatic stress disorder; SAD, social anxiety disorder; SI, suicidal ideation.

**Extended Data Table 2:** Recording duration and electrode implant locations of study participants.

| Subject | Total data length | Brain regions with implanted electrodes |
| --- | --- | --- |
| S1 | 667 days | Amygdala, hypothalamus, anterior limb of the internal capsule |
| S2 | 711 days | Subgenual cingulate, orbitofrontal cortex |
| S3 | 501 days | Subgenual cingulate |
| S4 | 560 days | Amygdala, hypothalamus, anterior limb of the internal capsule |
| S5 | 243 days | anterior limb of the internal capsule, thalamus |

### Mood cycle phase encodes clinical depression severity

We first asked whether long-term fluctuations in mood have underlying rhythmic structure at multiday timescales. Across participants, we identified prominent multiday mood cycles across a spectrum spanning periods of 2–34 days (Extended Data Fig.2a). Timescales of multidien mood cycles were significantly correlated across all participant pairs (Pearson r = 0.42–0.88, all p < 0.001; Extended Data Fig. 2b).

**Extended Data Figure 2:**
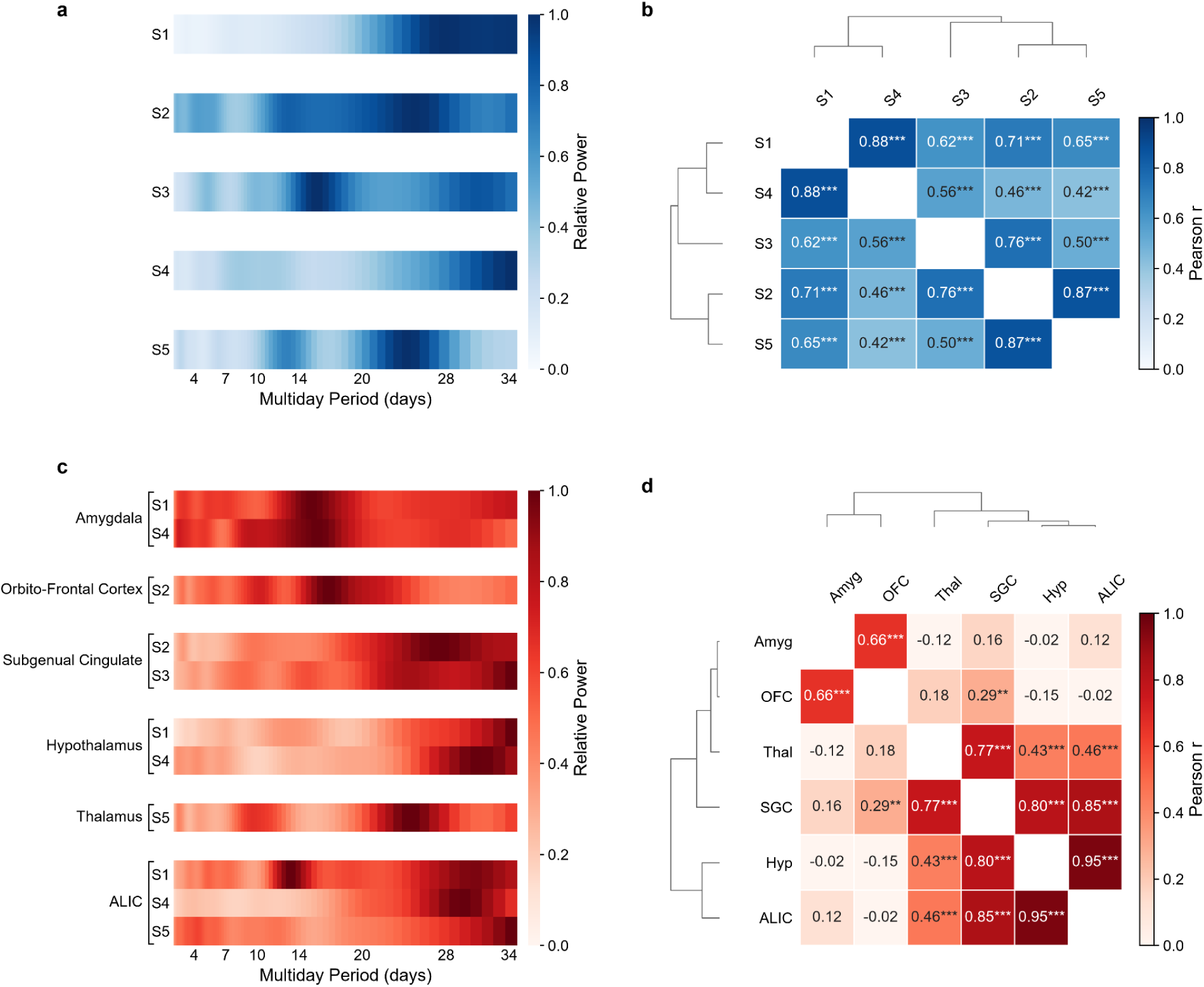
Multidien cycles organize neural activity and mood and are robust. **a.** Periodogram of VAS-D multidien cycle power per participant. **b.** Patient-by-patient Pearson correlation matrix comparing VAS-D periodograms. **c.** Periodogram of neural multidien rhythm power per participant and brain region (n=5 participants). **d.** Region-by-region Pearson correlation matrix comparing neural periodograms (*p < 0.05, **p < 0.01, ***p < 0.001)

We hypothesized that multiday cycling of mood is closely tied to depression severity. Specifically, momentary position within the mood cycle (also known as phase) may carry information related to clinical depression state. To measure momentary position in the mood cycle, we computed the instantaneous phase of the broadband multidien cycle in VAS-D for each participant. The broadband cycle summarizes the multidien activity as a weighted average over a range of timescales rather than relying on any single, narrow periodicity^16^, and has been used previously for seizure forecasting^17^. The instantaneous phase of this broadband signal indexes where the participant is in their mood cycle on a given day.

Dense sampling of VAS-D provided finer temporal resolution of the rising and falling portions of the mood cycle than the biweekly MADRS assessments. We therefore aligned both VAS-D ratings and MADRS scores to the phase of the broadband VAS-D cycle (Fig.2a). VAS-D varied systematically across the multidien mood cycle, with the highest ratings concentrated at the peak phase as shown in one representative participant S4 (Fig.2b; R = 0.17, p = 0.001, n = 560 days) and across participants (Fig.2d; R = 0.15, p = 0.001, cohort permutation test). MADRS scores showed a similar phase dependence to the peak phase of the mood cycle as shown in the same representative participant (Fig.2c; R = 0.27, *θ* = 179°, p = 0.009; n = 56 assessments) and across participants (Fig.2e; R = 0.26, *θ* = 125°, p = 0.010, cohort permutation test), indicating that the VAS-D-derived multidien cycle captured clinically meaningful variation in depression severity.

**Figure 2:**
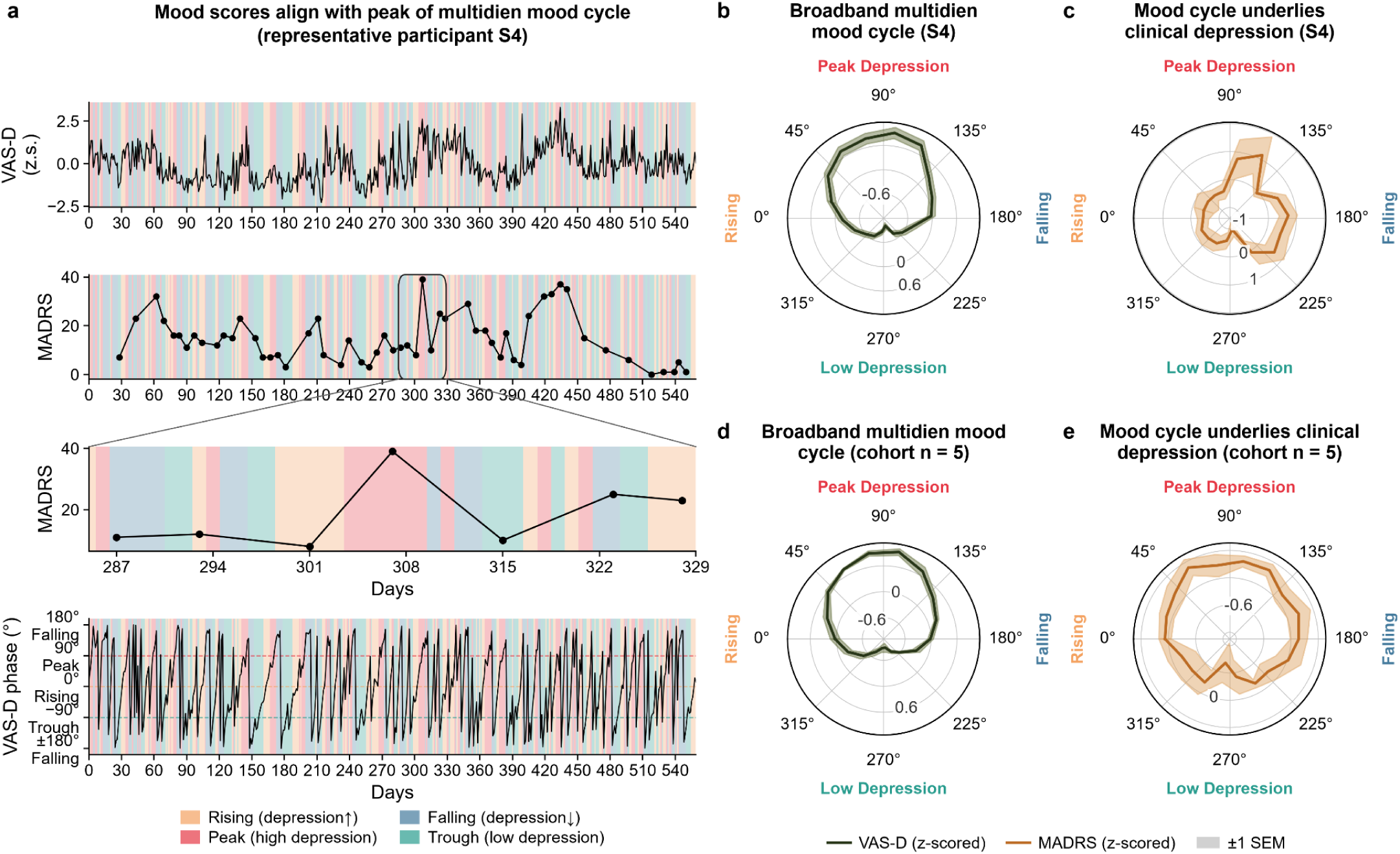
Clinical depression shows phase preference for multidien behavioral rhythms. **a.** Daily self-reported VAS-D (top), biweekly clinician-administered MADRS (middle, and zoomed-in), and the VAS-D phase of the broadband multidien rhythm (bottom) in one representative participant (S4). Background shading: VAS-D phase quadrants (yellow = rising, red = peak, blue = falling, green = trough). **b.** Mean VAS-D ± SEM (shaded) across broadband VAS-D phase (S4). Angular axis: phase (deg.); radial axis: mean VAS-D score (z-scored). VAS-D was concentrated near peak phase (R = 0.17, p = 0.001, n = 560 days). **c.** Mean MADRS ± SEM (shaded) across broadband VAS-D phase (S4). Angular axis: phase (deg.); radial axis: mean MADRS score (z-scored). MADRS was similarly concentrated near the peak phase of the mood cycle (R = 0.27, θ = 179°, p = 0.009; n = 56 assessments). **d.** Cohort VAS-D phase preference. Mean VAS-D ± SEM (shaded) across broadband VAS-D phase, averaged across all participants (N = 5). VAS-D was modulated by the multidien phase in every participant (mean within-participant R = 0.15, p = 0.001, 1000-permutation null - scores shuffled across phase, N = 5 participants). **e.** Cohort MADRS phase preference. Mean MADRS ± SEM (shaded) across broadband VAS-D phase, averaged across participants. MADRS was phase-modulated at the cohort level (mean within-participant R = 0.26, θ = 125°, p = 0.010, 1000-permutation null - scores shuffled across phase, N = 5 participants).

Our results suggest that the phase of the mood cycle serves as a compact descriptor of where a participant lies within an evolving depression trajectory, including whether symptoms are increasing, decreasing, near a peak, or near a trough. Phase-based representation of an individual’s current position and their direction in the depression trajectory (which cannot be obtained from the depression score alone) provides a clinically interpretable tool for estimating time-varying mood symptom severity.

To establish that mood cycles describe the trajectory of depression symptom fluctuations, we trained Ridge regression models to predict future VAS-D scores. We separately evaluated the predictive performance of three sets of features: multiday mood cycle amplitude (Fig.3a), multiday mood cycle phase (Fig.3b), and the raw VAS-D score (Fig.3c), and evaluated forecasting performance across different time horizons (e.g. H = 1 implies 1 day ahead). Phase-based forecasting was significant in every participant and achieved a mean held-out test correlation of r = 0.77 (SD 0.13), range 0.58–0.93 (p < 0.001) for next-day (H = 1) VAS-D prediction (Fig. 3b). Phase features showed a trend over amplitude and raw VAS-D score (Fig.3d). Phase-based forecasts remained significantly above the permuted-target null across horizons up to 30 d (p<0.05; Fig.3e), while amplitude and score forecasts remained significant up to 14- and 7-day horizons respectively. Sensitivity analysis, performed by varying the fraction of training data (0.50–0.75) and measuring the predictive performance (Pearson’s r), showed phase exceeding the permuted-null across all fractions (p < 0.05; Fig.3f). The stability of the phase-based model across training fractions indicates that cycling information is robust across different training-set sizes. Phase-based models exposed to progressively less training data consistently outperformed models based on amplitude and raw score and yielded reliable mood forecasts.

**Figure 3:**
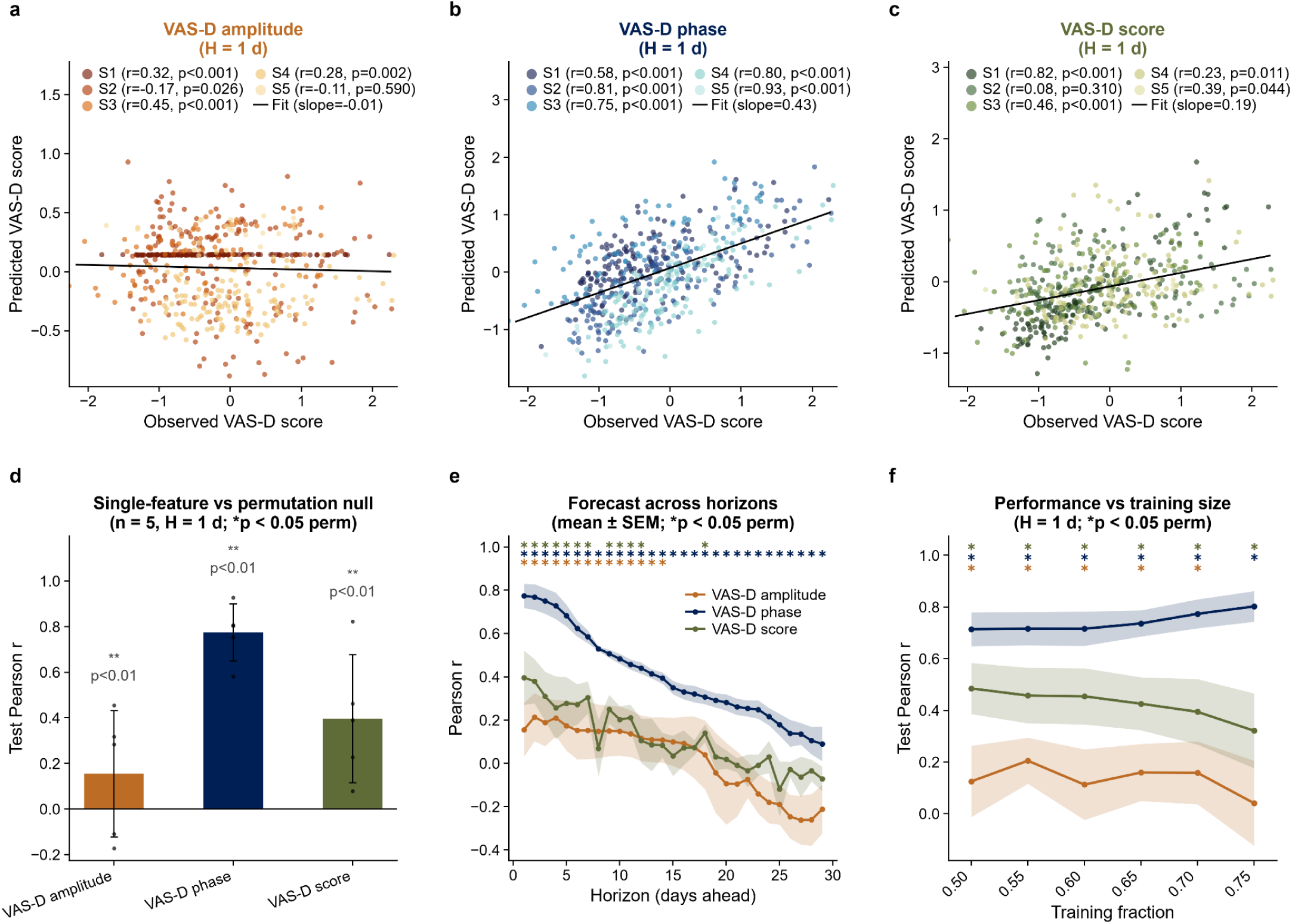
Phase of multiday mood rhythm explains depression trajectory. **a–c.** Observed versus predicted next-day VAS-D (forecast horizon H = 1 d) from RidgeCV models trained on the VAS-D multidien amplitude (a), phase (b), or the raw VAS-D score (c), across all wavelet scales (2-34 days). **d.** Test-set Pearson r per predictor at H = 1 d (bars, mean ± SD; dots, individual participants); asterisks, predictors for which the cohort-mean Pearson r exceeds the shuffled-target null (p < 0.05, 100 permutations). **e.** Forecast accuracy (Pearson r, mean ± SEM) across forecast horizons (1-30 d). Asterisks: horizons at which the cohort mean exceeds the shuffled-target (p < 0.05, 100 permutations). **f.** Forecast accuracy (Pearson r, mean ± SEM) as a function of training fraction (0.50–0.75, H = 1 d). Asterisks as in e.

Together, our results suggest that phase integrates information across multiple time horizons into an instantaneous feature that provides it further reach than amplitude or raw score for forecasting. While raw mood score primarily describes a participant’s current depression level, phase specifies position within the mood cycle and therefore whether mood is moving toward higher or lower severity. This raises the possibility of using the mood cycle phase as an anticipatory index of heightened mood symptom severity.

### Neural rhythms pace mood cycles

We next examined whether neural activity exhibits multidien rhythms that occur in lockstep with mood cycles. Across participants, we observed multiday rhythms in the iEEG signal spanning periods of 2–34 days (Extended Data Fig.2c). Timescales of multiday rhythms were significantly correlated in 8 of 15 region pairs (Pearson r = 0.29–0.95, all p < 0.05; Extended Data Fig. 2d), revealing two clusters of brain regions with similar multidien periodicities. Subcortical and midline regions, which included SGC, hypothalamus, ALIC, and thalamus, formed one tightly correlated cluster (r = 0.43–0.95). Amygdala and OFC formed a second correlated cluster (r = 0.66) and showed weak, non-significant correlations with the subcortical–midline cluster (r = −0.15–0.18). These results indicate that neural multiday rhythms are a prevalent and heterogeneous feature in depression.

We next asked whether neural multiday rhythms are synchronized with multiday mood cycles. We used coherence, a measure of phase alignment between two oscillatory signals whose value ranges between 0 (no synchrony) and 1 (complete synchrony), to quantify synchronization between iEEG rhythms and mood cycles across multiday periods (Fig.4a). We found that iEEG signals and VAS-D scores showed coherence across a wide spectrum of multiday timescales (Fig.4b), both within and across participants across brain regions (Extended Data Fig.3a). Mood–neural coherence was observed in every sampled region and in all five participants, despite differing implant locations. Peak coherence periods did not differ significantly across regions (Kruskal–Wallis H = 4.19, p = 0.523), though sample sizes per region were small (n = 1–3). Additionally, coherence was positively correlated across all region-pairs (Pearson r = 0.26–0.88, all p < 0.05; Extended Data Fig. 3b). Multidien mood–neural coupling therefore appears distributed across the cortico-limbic network rather than confined to a single structure.

**Figure 4:**
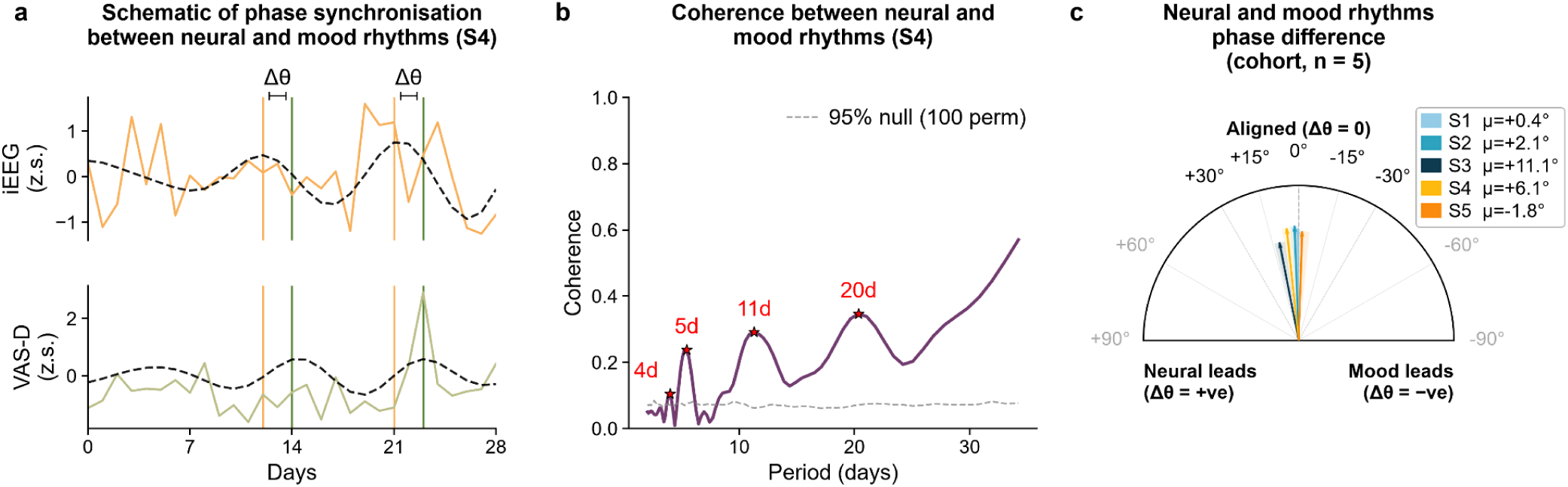
Multidien rhythms in iEEG and behavior are synchronized, with neural rhythms leading. **a.** Daily iEEG activity in one example limbic iEEG channel (amygdala) (top, orange) and VAS-D (bottom, olive) over a representative 28-day window in one representative participant (S4); black dashed lines: wavelet reconstruction; Δθ: phase difference between iEEG cycle and VAS-D cycle. **b.** Coherogram showing coherence between iEEG and VAS-D multidien rhythms; asterisks: coherence peaks at 5, 11, and 20 d. Dashed line: 95% confidence interval (100 permutations). **c.** Phase difference between iEEG rhythms and mood rhythms across all recording channels for each participant (n=5 participants). Δθ = 0° (top) indicates aligned cycles, positive Δθ (left) indicates neural-leading, negative Δθ (right) indicates mood-leading. Arrows: per-participant circular-mean phase difference pooled across the four iEEG channels and time; shaded wedges: 95% bootstrap CI (3000 resamples); arrow length: mean resultant vector R; per-participant μ in legend. Four of five participants show μ > 0, consistent with a neural-leading trend.

**Extended Data Figure 3:**
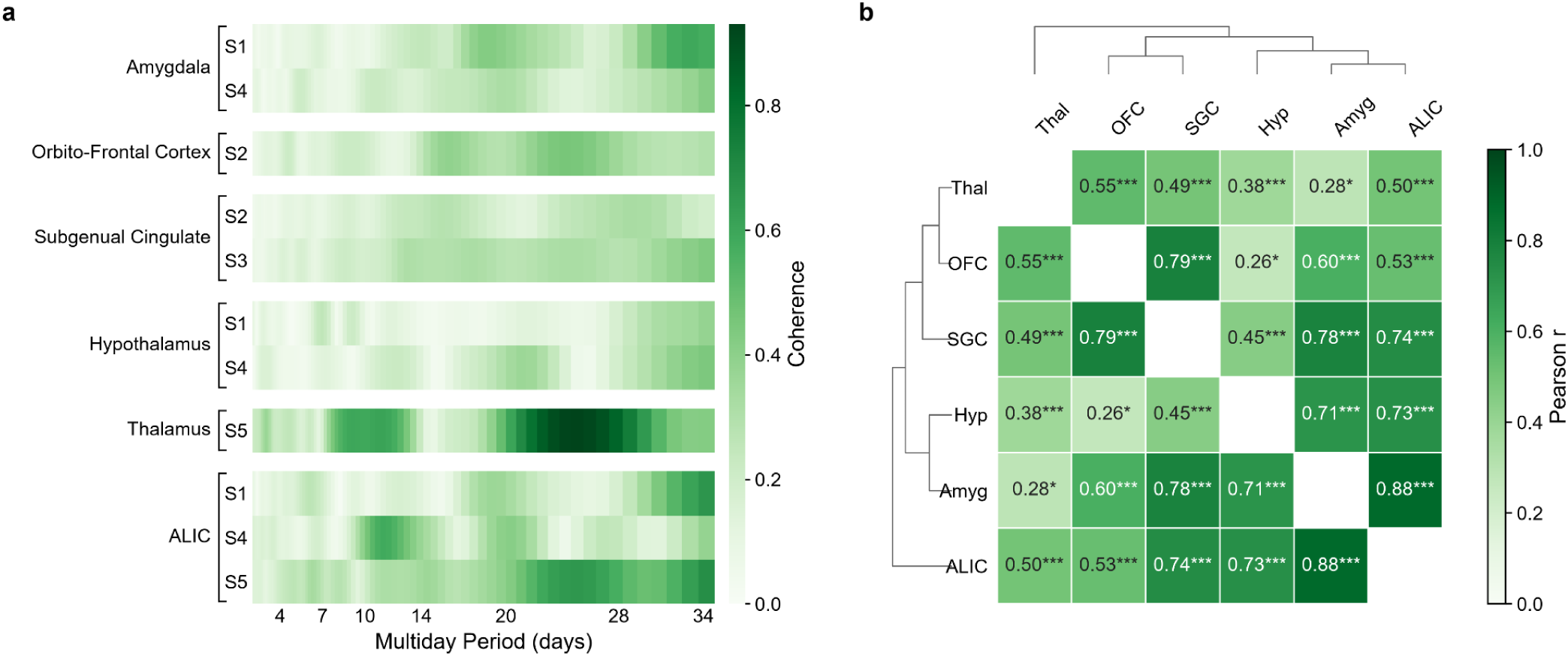
Neural and mood multidien cycles are synchronized. **a.** Coherogram between neural and mood multidien cycles across participants (n=5). **b.** Region-by-region Pearson correlation matrix comparing coherence spectral profiles across brain regions.

Coherence also captured underlying mood-iEEG dynamics that were masked by conventional static correlation. In a representative participant (S4), correlation between iEEG activity and VAS-D computed over the whole recording period was negligible (r = −0.03, p = 0.55; Extended Data Fig.4a), and yet, a 14-day rolling correlation between the two signals showed high-correlation epochs recurring at 5–7, 10–11, and 18–20-day periods, which matched the peak periodicities in the coherence between the two signals (p<0.05, Extended Data Fig.4b–c). We replicated this phenomenon in two simulated coherent rhythms (Extended Data Fig. 4d–f), indicating that the neural-mood relationship is fundamentally one of multiday phase coherence that may be poorly resolved by a conventional static correlation approach. This highlights the importance of coherence as a metric for understanding time-varying brain-behavior relationships.

**Extended Data Figure 4:**
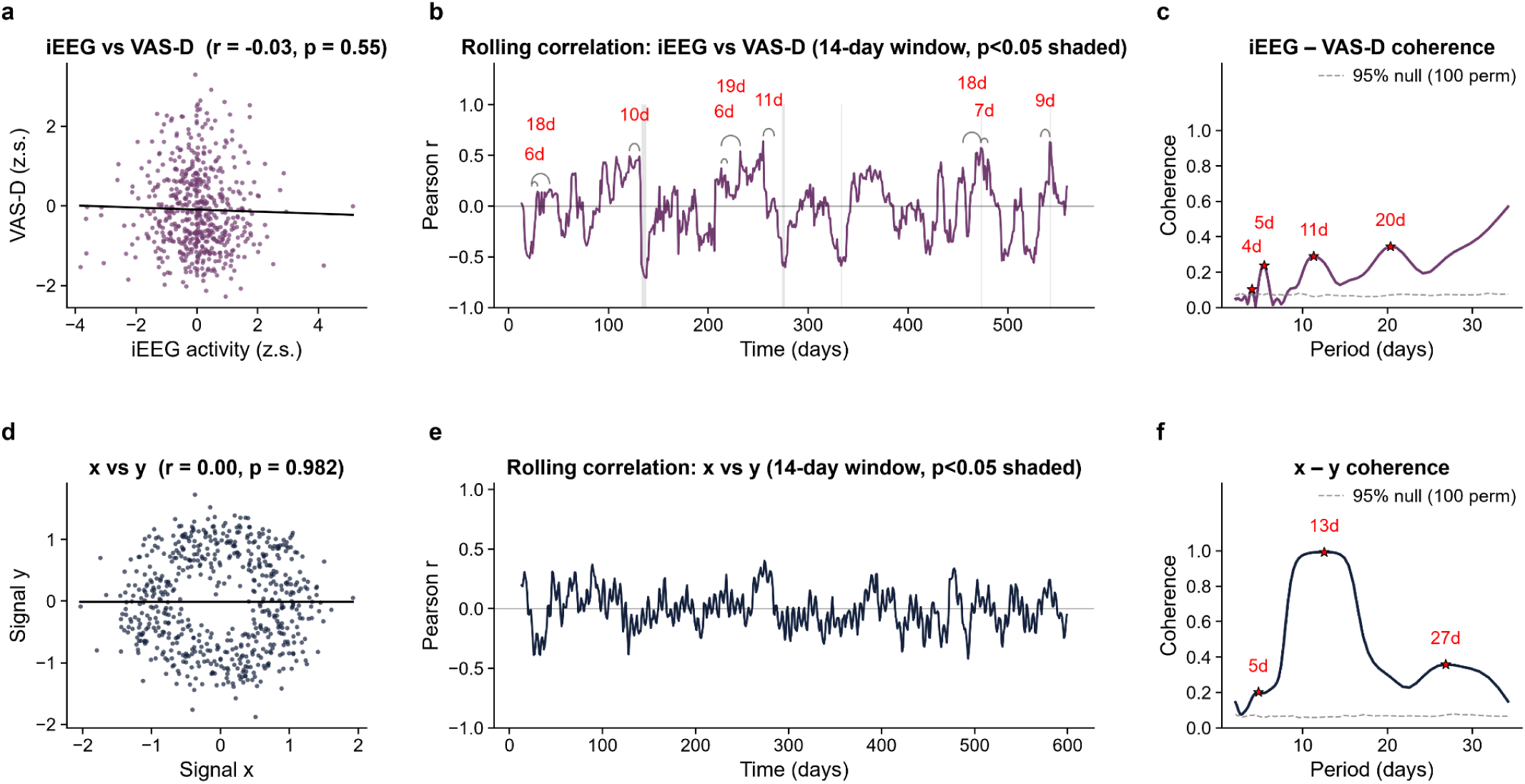
Conventional correlation masks underlying synchronisation of iEEG and mood. Top row, representative participant S4 (real data); bottom row, simulated data. **a.** Daily iEEG activity (representative limbic channel, amygdala) plotted against VAS-D depression score (r = −0.03, p = 0.55). **b.** Pearson correlation between the two signals computed in a 14-day rolling window; grey shading: windows in which the correlation is significant (p < 0.05). The correlation swings between positive and negative values, and high-correlation epochs (arcs, labelled with their separation in days) recur at intervals close to the significant coherence periods identified in (c). **c.** Coherence between iEEG and VAS-D, computed from their wavelet transforms, across multiday periods (2-34 days). Dashed line: 95th percentile of null distribution (VAS-D shuffled, 100 permutations); asterisks: coherence peaks exceeding null (∼4-6, 10-11 and 18-20 days). **d-f.** The same three analyses applied to two simulated sinusoids (signal x, signal y) of period 12 days separated by a constant 90° phase offset (signals are coherent by construction), plus Gaussian noise. **d.** Conventional static correlation between x and y (r = 0.00, p = 0.982). **e.**14-day rolling correlation. **f.** Coherence between x and y with a dominant peak at the simulated 12-day period (red asterisk). Panels d-f show that a static correlation collapses the neural-mood relationship to near zero even when the two signals are strongly coherent, and that the fluctuations seen in windowed correlations indicate the presence of this coherence.

Additionally, we evaluated if mood cycles and neural rhythms show temporal ordering with one leading the other. We computed the per-day phase difference between the broadband iEEG rhythm and mood cycle, pooled across regions sampled per participant (Fig.4c). In four of five participants, neural rhythms consistently preceded (led) mood cycles, corresponding to a positive mean phase difference (μ) between the two signals (p<0.05 in S2, S3, S4; same directional trend present but did not reach significance in S1). The fifth participant (S5) did not exhibit a statistically significant effect of neural rhythms leading mood cycles, potentially driven by limited statistical power due to short data duration (243 days). No participant showed a significant mood-leading phase difference, supporting a consistent neural-leading pattern across the cohort where the effect is detectable.

Together, these observations indicate that iEEG multidien rhythms are phase-locked to mood cycles and lead mood cycles. This finding suggests the possibility of using iEEG rhythms as both an indicator of future mood, as well as an intervention target to modify future mood, even before it starts to show indications of worsening.

### Neural multidien phase forecasts the mood trajectory

Based on the lead-lag relationship between neural rhythms and mood cycles, we constructed a forecasting model using the multiday iEEG rhythm to predict depression trajectory.

Specifically, we trained a Ridge regression model to predict the future phase of the multidien mood cycle using neural multidien rhythm phase (Fig. 5a–c) and evaluated forecasting performance across different time horizons (e.g. H = 1 implies 1 day ahead). For each participant, the multidien neural phase from all four iEEG channels recorded by the RNS System were inputted as the model predictor. Phase prediction accuracy was quantified as the mean resultant vector R of the phase differences between predicted and observed VAS-D phase, where higher value reflects more consistent phase tracking; significance was assessed against a permutation null obtained by shuffling observed phases. All five participants showed significant phase prediction accuracy averaged across forecast horizons (mean R over H > 1; permutation p < 0.05; Fig.5d, Extended Data Fig.5). In every participant the phase model exceeded the accuracy of baseline models trained on same-day raw iEEG voltage, VAS-D score history, or their combination. At the cohort level, mean phase prediction accuracy of the phase model remained significant at every individual forecast horizon from 2 to 30 days (mean ± SEM across participants; permutation p < 0.05; Fig. 5e), and consistently above all baseline models. To assess whether forecasting depended on specific recording sites, we systematically varied which iEEG channels were included in the model, training it on every possible combination of one, two, three, or four channels (15 combinations total) for each participant. Single-channel models reached significance in 20 of 20 participant-channel combinations, and accuracy did not differ significantly with additional channels (Friedman test, χ² = 3.48, p = 0.32, n = 5; Extended Data Fig.6). This indicates that the multidien phase-based forecasting ability is distributed across the sampled cortico-limbic network and not limited to any single contact.

**Figure 5:**
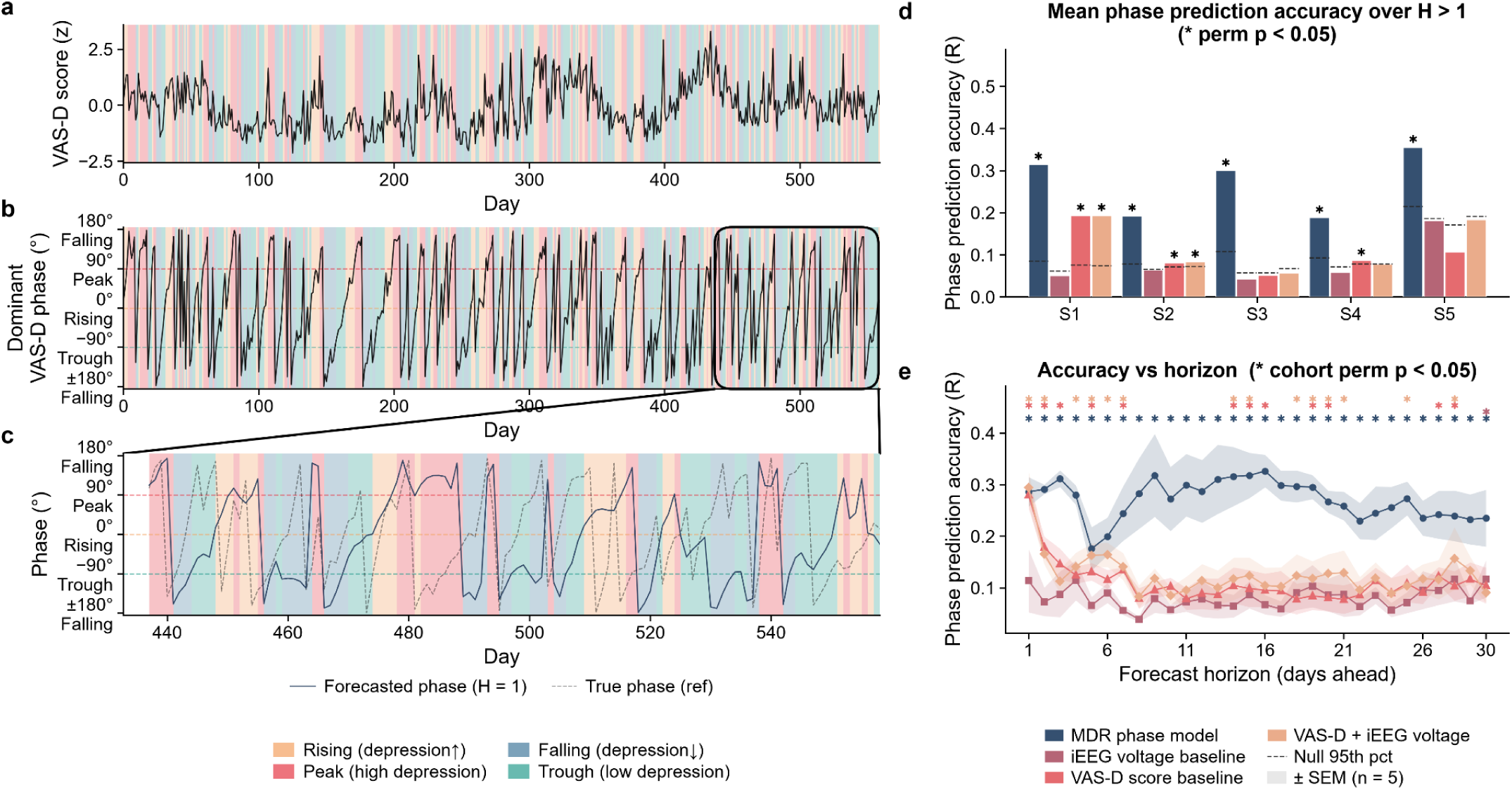
Neural multidien phase forecasts depression cycle phase. **a.** VAS-D score over time for S4, color-coded by VAS-D multidien cycle phase (yellow = rising, red = peak, blue = falling, green = trough). **b.** Broadband VAS-D multidien phase over time for S4, color-coded as in (a). **c.** Inset of the test period (boxed in b) showing forecasted (solid, H = 1 day) versus true (dashed) broadband VAS-D phase for S4. **d.** Phase prediction accuracy for each participant (calculated as the mean resultant vector R of the phase difference between the observed and neural-phase-forecasted VAS-D phase), across all forecast horizons H = 2–30 days. Colored bars: multidien-phase model (MDR), iEEG baseline model, VAS-D score model, VAS-D + iEEG model. Higher R value indicates more consistent phase tracking. Dashed line: 95th percentile of the horizon-averaged permuted null. Asterisks: R exceeds null (p < 0.05). **e.** Phase prediction accuracy (mean ± SEM across participants, N = 5) as a function of forecast horizon (1-30 days). Shaded bands: ± SEM. Asterisks: horizons at which cohort-level accuracy significantly exceeds the permutation null (p < 0.05); each row corresponds to one model, with the cohort test computed as the mean within-participant R tested against the averaged per-participant permutation nulls.

**Extended Data Figure 5:**
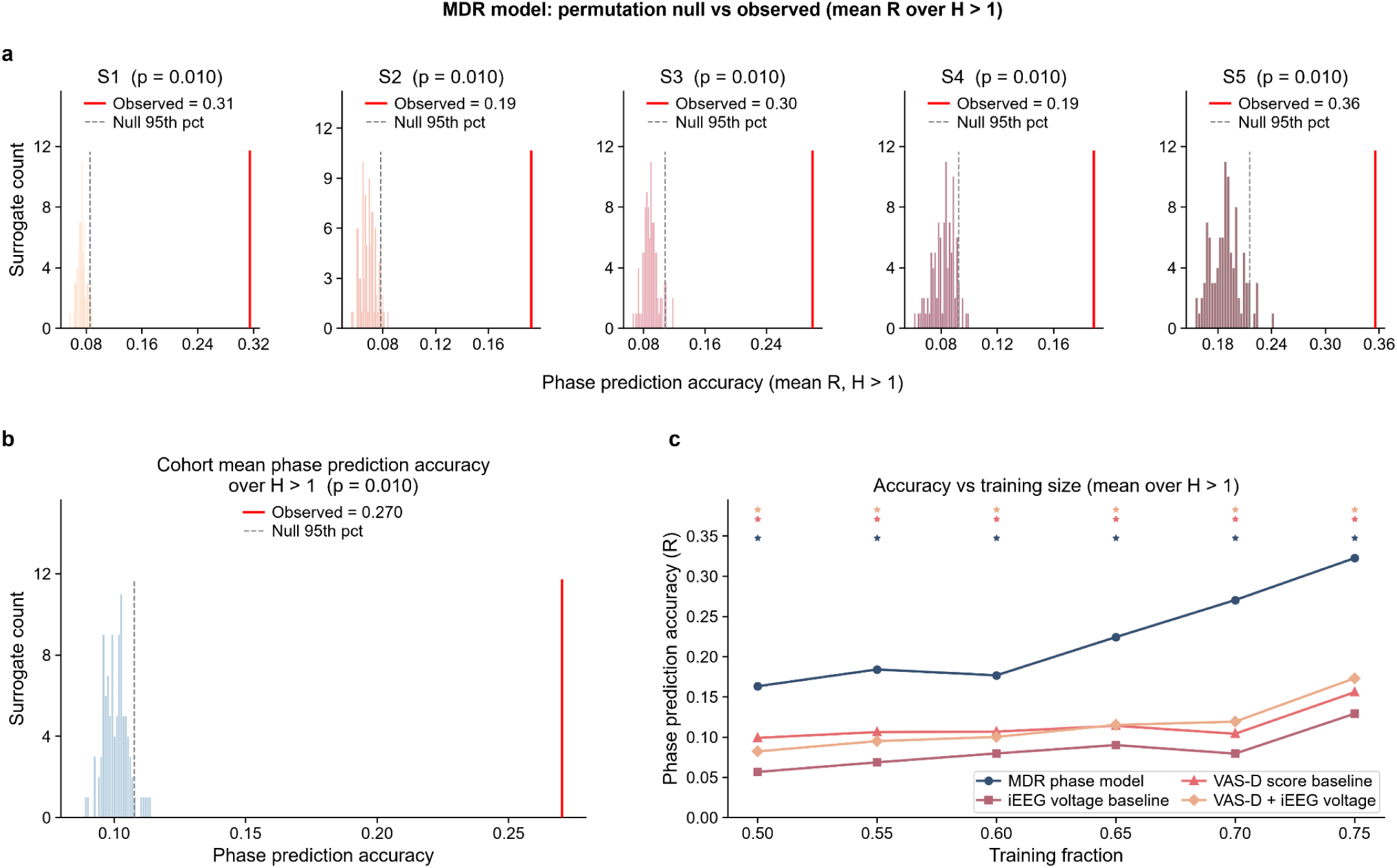
Neural multidien phase forecasts mood multidien cycle. **a.** Per-participant permutation null distributions of mean phase prediction accuracy (R averaged over H = 2–30 days) for the MDR phase model (S1-S5). Surrogate null obtained by shuffling the observed phases and averaging surrogate R across horizons; red line: observed mean phase prediction accuracy; dashed line: null 95th percentile. **b.** Across the cohort (n = 5), accuracy exceeded chance (mean R = 0.270, permutation p = 0.010. **c.** Phase prediction accuracy (mean R over H = 2–30) as a function of the fraction of data used for training (0.50–0.75); asterisks: permutation p < 0.05. Accuracy was sustained across training fractions, indicating robustness to reductions in training-set size.

**Extended Data Figure 6:**
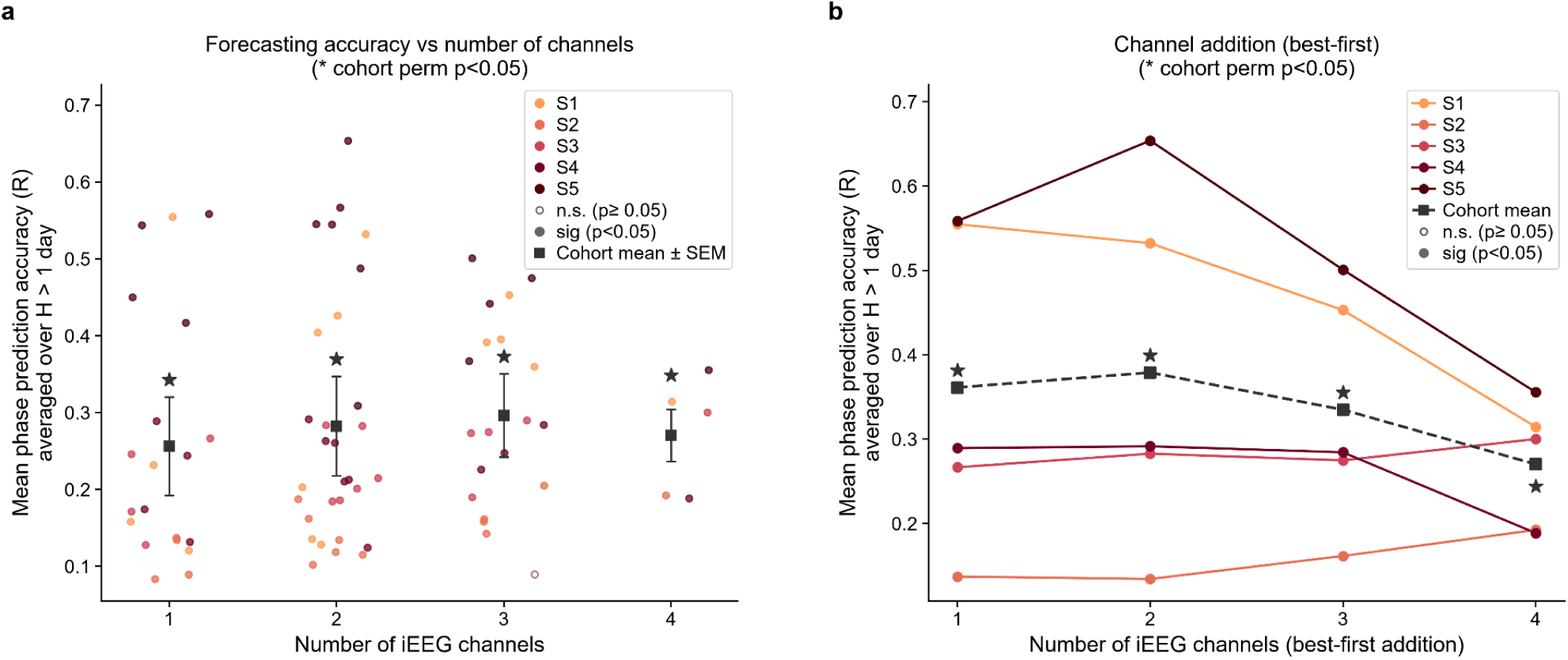
Forecasting accuracy is distributed across limbic recording sites. **a.** Mean phase prediction accuracy (R, averaged over H = 2–30 days) as a function of the number of iEEG channels included in the MDR phase model. Each dot represents one participant × channel-subset combination (4 single-channel, 6 two-channel, 4 three-channel, and 1 four-channel subset per participant). Filled dots indicate significance (permutation p < 0.05); hollow dots indicate non-significance. Black squares: cohort mean ± SEM across participants (asterisks: cohort permutation p < 0.05). Single-channel models reached significance (permutation p < 0.05) in 20 of 20 participant-channel combinations, indicating that the multidien forecasting signal is accessible from any sampled limbic contact. Accuracy did not differ significantly across channel counts (Friedman test, χ² = 3.48, p = 0.32, n = 5). **b.** Mean phase prediction accuracy as a function of the number of channels added to the single channel showing highest R for each participant. At each step, the channel yielding the highest mean R when added to the current set was selected. Filled and hollow markers denote significance as in (a); asterisks indicate cohort permutation p < 0.05. Cohort-mean accuracy (dashed black) peaked at two channels and declined with additional channels, suggesting redundant phase information across contacts and increasing feature dimensionality relative to training-set size.

Together, these results indicate that the instantaneous phase of the iEEG multidien rhythm carries forward-looking information about the participant’s mood trajectory days to weeks in advance.

### Electrical stimulation modulates multidien rhythm dynamics

Based on our finding that multidien rhythms pace the severity of mood symptoms, we asked whether chronic neurostimulation modulates these rhythms. Using double-blinded sham-controlled neurostimulation conducted as a part of the parent PRESIDIO clinical trial, we evaluated stimulation effects on multidien neural rhythms across three conditions. In the closed-loop stimulation (CL) condition, the device monitors a neural biomarker that signals when a participant is in a high-symptom state and stimulates only at those moments. In the intermittent stimulation (INT) condition, stimulation is delivered on a fixed schedule unrelated to the biomarker. In the sham condition no stimulation is delivered. Across these three conditions, we examined how stimulation effects on multidien rhythms depend on stimulation timed to the high-symptom state (CL), or on interval-based stimulation regardless of timing (INT).

We hypothesized that stimulation would modulate the strength of these rhythms and potentially elongate the timescale of these rhythms so that peak depression occurs more rarely. To test this hypothesis, we evaluated two metrics of stimulation effect: (i) amplitude (power) of the multidien rhythm, which indexes how strongly the cycle is present, and (ii) phase distribution across the cycle, which indexes how individuals move between high and low depression.

Of the 5 participants included in this analysis, only three had undergone a blinded cross-over comparison across stimulation conditions. We compared their multidien rhythm properties across days when stimulation was kept on (CL, INT) in comparison to when it was kept off (sham). We observed a shift in the distribution of multidien rhythms towards slower periods, as well as an increase in rhythm amplitude, in stimulation periods compared to sham in one representative participant (Fig. 6a) and across participants (Extended Data Fig.7). Compared to sham, broadband multidien rhythm amplitude significantly increased in closed-loop stimulation periods (p<0.035) and in intermittent stimulation periods (p<0.001) across participants (Fig.6b). Stimulation also reshaped the durations of the multidien rhythm phases: the fraction of days spent in the peak phase reduced, while the fraction of days spent in the transitory falling and rising phases increased (Fig.6c; p = 0.002).

**Figure 6:**
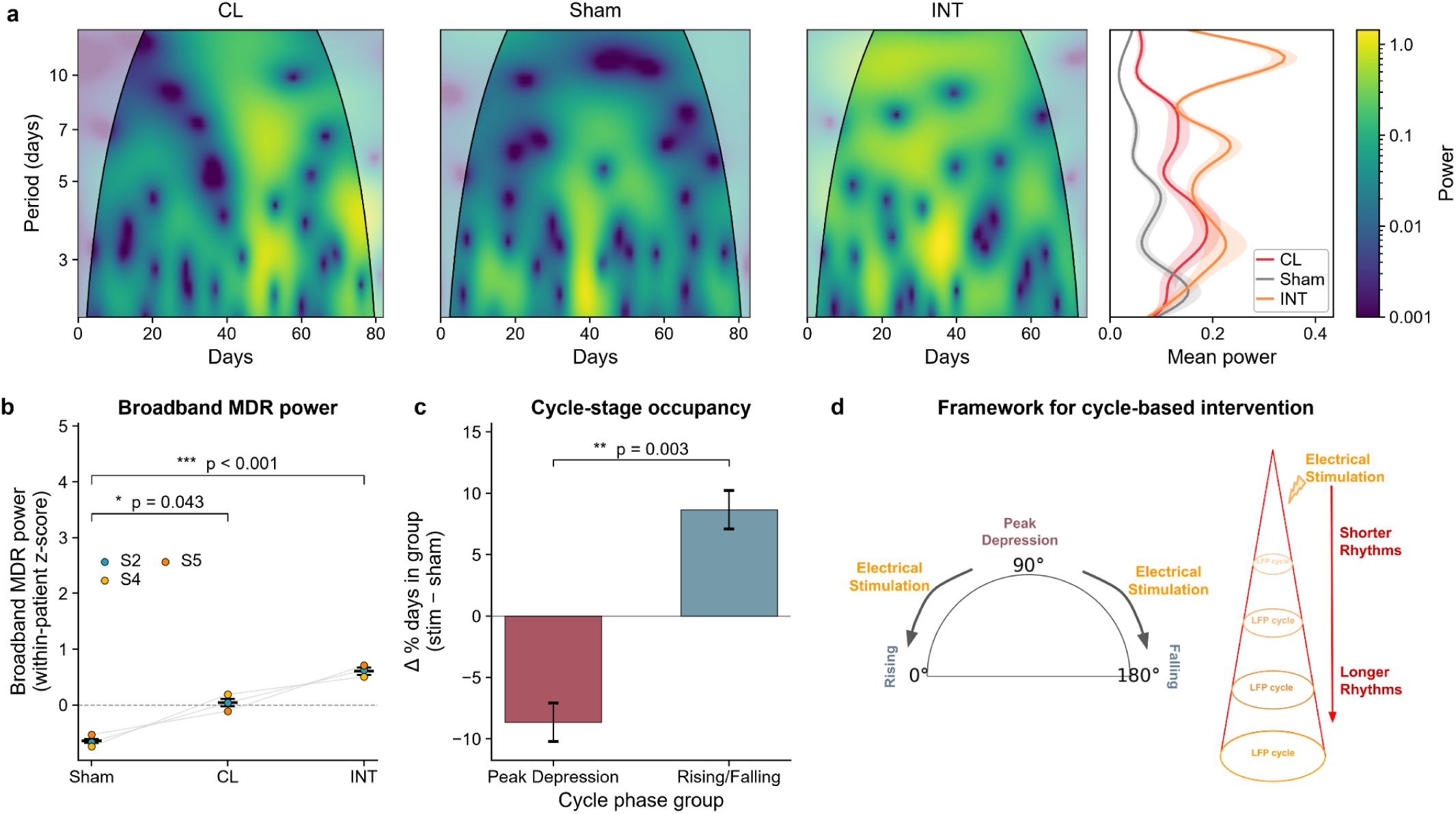
Electrical stimulation modulates neural multidien rhythms. **a.** Spectrogram showing multidien wavelet power across conditions (CL = closed-loop stimulation, Sham = device turned off, no stimulation, INT = intermittent stimulation not based on biomarker detection) in one representative participant (S5) and corresponding periodogram showing power averaged over time (ordering of the conditions has been shuffled to preserve anonymity). **b.** Broadband multidien cycle power in each condition across participants (n=3). **c.** Stimulation-induced change in percentage of days in peak depression phase vs. rising/fall phase of the neural multidien cycle (n = 6; CL, INT x 3 participants, mean ± SEM). **d.** Schematic model of the effect of stimulation on multidien cycles: Stimulation slows down the multidien iEEG rhythm and thereby pushes participants away from the peak depression phase.

**Extended Data Figure 7:**
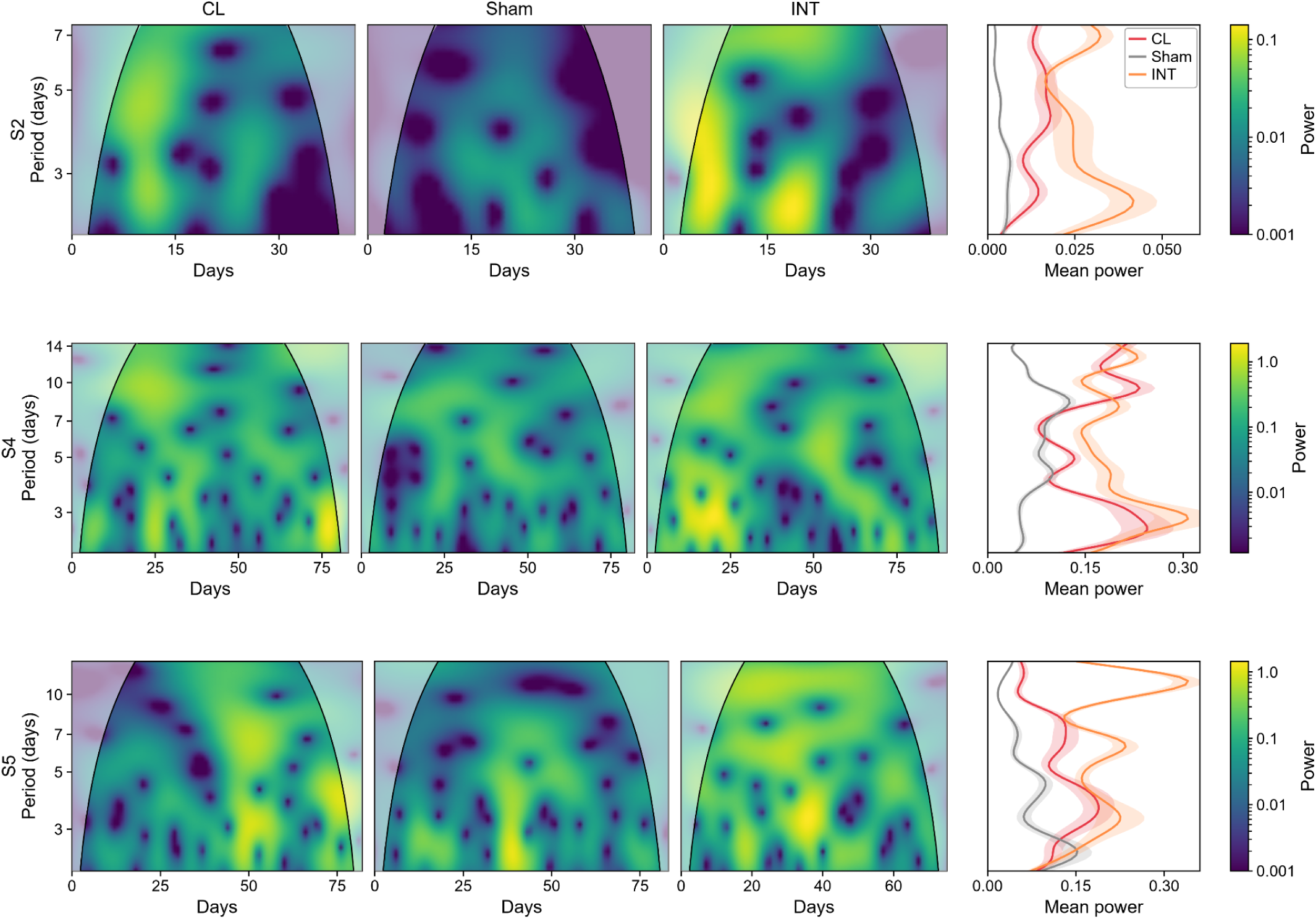
Stimulation modulates neural multidien cycles. Spectrogram showing multidien wavelet power across conditions (CL = closed-loop stimulation, Sham = device turned off, no stimulation, INT = intermittent stimulation not based on biomarker detection) across participants (n=3).

Together, these results show that electrical stimulation increases broadband multidien rhythm period and amplitude and redistributes phase occupancy, reducing the fraction of days spent in the peak-symptom phase (Fig.6d). These findings indicate that neurostimulation can modify the temporal structure of multiday neural rhythms, offering a potential mechanism for reducing time spent in states of elevated mood symptom severity.

## Discussion

In summary, using years-long brain recordings in adults with severe major depressive disorder undergoing responsive neurostimulation treatment, we reveal multidien rhythmic cycling in limbic network activity and mood-related symptoms. Clinical depression severity rising at peak phase of the underlying mood cycle suggests the cycle acts as a temporal manifold where the cycle phase encodes not only the participants’ current position, but also their direction in the depression trajectory. This enables estimation of mood symptom as a practical clinical endpoint. Multidien iEEG rhythms and mood cycles were synchronized – with multidien rhythms in limbic network activity predicting mood cycle trajectory up to 30 days in advance. Narrow phases of these rhythms were tightly linked with fluctuation in depression severity, suggesting that depression is organized by brain network activity operating over multiple timescales that modulate disease state. Additionally, closed-loop and fixed-interval intermittent electrical stimulation reshapes multidien rhythm dynamics and shifts time away from peak depression phases of the cycle. Together, these findings establish multidien rhythms as a previously unrecognized organizing principle of pathological brain-behavior dynamics in depression, offering a tractable temporal substrate for next-generation chronotherapeutic neuromodulation.

Our findings extend a multidien rhythm framework of multimodal physiological signals^6–9,17,18^, first developed in epilepsy for gauging the risk of seizure occurrence^6–8^, into psychiatry, helping to explain the temporal dynamics of mood disorders. Three points of convergence emerge. First, the dominant cycle lengths we observe in MDD overlap closely with those reported in epilepsy^6^, suggesting a shared timescale of slow neural organization across diverse pathologies. Second, as in epilepsy^6^, peak symptom severity is paced by an underlying rhythm of brain network activity. Additionally, the robustness of our findings across participants, and the fact that multidien cycles have been shown previously in depression^10^, bipolar disorder^11^, and in healthy human heart rate^4^ and smartphone usage^5^, and slow changes in neural dynamics have been shown to track depression recovery^12,13^, collectively suggest that slow cyclical organization of neural activity and behavior is a general biological principle across neurological and psychiatric disorders. Our work provides a direct human neural correlate for this long-suspected slow temporal architecture of mood, and a framework for operationalizing neural timescales to forecast mood in depression. The observance of this temporal architecture directly in iEEG argues these slow multidien rhythms are an organizational principle akin to traditional power spectra bands.

A central finding in our study is that multidien rhythms in iEEG are synchronized with and lead multidien mood cycles across participants. Prior studies have elegantly demonstrated the ability to decode instantaneous mood states from contemporaneous limbic neural activity^19,20^. Our study leverages chronic recordings to predict mood days ahead. Our results also align with recent demonstration that subcallosal cingulate dynamics trace depression recovery and predict relapse in participants with depression^12,13^, and that disruption of ventral striatal rhythmicity predicts response to deep brain stimulation (DBS) in participants with obsessive-compulsive disorder (OCD)^21^. Whereas these studies established that slow neural dynamics track stimulation-induced clinical change, our findings reveal an endogenous cyclical architecture that organizes mood independently of stimulation, and can forecast symptom trajectory days to weeks ahead.

Several features of the multidien biomarker we describe distinguish it from prior slow-dynamic signatures of depression. First, our results show that a momentary assessment of mood state (daily VAS-D score) is a comprehensive marker of both immediate short-term distress and long-term clinical depression (as assayed by MADRS), which occur at different timescales. This suggests multidien mood cycles index information across timescales. Second, we report presence of the multidien biomarker across different limbic structures, complementing prior work which found long-term biomarkers of depression in subgenual cingulate^12,13^. This indicates this slow temporal architecture may be a distributed property of the limbic network rather than a feature of any single node. Third, the multidien biomarker we report encodes continuous ongoing mood fluctuations as opposed to discrete clinical transitions like recovery and relapse. For closed-loop neuromodulation applications, a continuous readout of symptoms offers an advantage over a biomarker of relapse as it enables proactive phase-timed interventions rather than reactive response to relapse.

Finally, we observed the multidien biomarker across both stimulation and non-stimulation periods, suggesting it reflects endogenous organization rather than a therapy-induced signature. Our results point to a two-part mechanism by which stimulation acts on the neural multidien rhythm. First, stimulation strengthens rather than abolishes the rhythm. The rise in broadband amplitude under both active conditions compared to sham suggests stimulation more strongly recruits the limbic network underlying the rhythm. Second, and more consequentially, stimulation rebalances an individual’s phase occupancy across the rhythm, shortening the high-symptom peak phase and lengthening the lower-symptom rising and falling phases. Thus participants spend fewer days in their most severe depressive state and more days in milder ones. This indicates that stimulation modulates these multidien rhythms. Our findings indicate that the effect of stimulation of multidien rhythms might be chronotherapeutic rather than acutely suppressive - instead of erasing the rhythm, stimulation reshapes its temporal architecture by compressing the window of greatest symptom burden. This raises the possibility that timing stimulation relative to multidien cycle phase might be a critical and underappreciated determinant of clinical response. This mechanism motivates cycle-informed adaptive stimulation timed to an individual’s multidien phase, forming the basis of a chronotherapeutic approach to neuromodulation.

Our findings suggest ultra-slow neural oscillations modulate an emerging concept of *depression risk*, which we define as a time-varying increase in susceptibility to external events, internal stressors, or stimuli that might otherwise be tolerable, leading to worsening depressive symptoms. We present depression risk as a quantifiable property of an individual’s multidien neural and mood dynamics. Our results indicate that the phase of the multidien mood cycle indexes an individual’s momentary position along a recurring depression trajectory. First, we found depressive symptom severity to be concentrated at the peak of the mood cycle, suggesting depression risk is not uniform across the cycle and is localized to identifiable phases. Second, the multidien neural rhythm led and forecasted the mood cycle days to weeks in advance, indicating elevated depression risk can be anticipated before symptoms intensify rather than in retrospect. Third, electrical stimulation biased participants away from the high-symptom phases, indicating mood risk is a modifiable target. This depression risk framework thus presents a dynamic view of depression in which a person continuously traverses windows of heightened and diminished vulnerability, and in which intervention timing relative to the cycle becomes important.

While prior studies have demonstrated the importance of neuromodulation for depression, timing stimulation for maximum positive effect remains an open challenge that could be addressed with the multidien rhythm framework. Deep brain stimulation of the subgenual cingulate produced response in seminal work^22^ but failed to separate from sham in the subsequent BROADEN trial^23^. Intracranial studies have demonstrated that responses to stimulation depend on the participant’s symptom state at the time of delivery^14,24^, motivating the search for state-aware biomarkers. However, temporal instability in brain-behavior relationships has emerged as a central bottleneck for psychiatric neuromodulation paradigms that rely on a stable biomarker. Our results suggest that this temporal instability might be due to an underlying slow oscillatory architecture. Correlation between limbic network activity and mood systematically varied over time and traversed a low-dimensional cyclical manifold whose phase carried predictive information that instantaneous correlation does not. Multidien phase-based biomarkers can thus serve a dual role: forecasting of symptom severity days to weeks ahead, and serving as an intervention target for preventative treatment.

Mechanistically, we speculate that the drivers of multidien rhythms in mood may be similar to systems implicated in epilepsy^6,9^. Possible sources include changes in physiological systems (temperature, heart rate, accelerometry and electrodermal activity)^9^, sleep-wake cycle^25^, and endocrine systems (about-weekly rhythms in melatonin, aldosterone, growth hormone and cortisol)^26^, which is supported by findings such as hypothalamic-pituitary-adrenal axis dysregulation in depression^27,28^. The neural-leads-behavior directionality we observe suggests limbic circuit dynamics may be the substrate through which these slower drivers shape symptom expression, although mutual entrainment by a common upstream driver cannot be excluded by our correlational design. Dynamical-systems models of depression have proposed oscillatory dynamics underlying mood^29^, which might then be entrained by environmental Zeitgebers (‘time-givers,’ such as lunar tidal cycles in bipolar disorder^11^). We thus speculate that the multiday rhythms we report might be generated and shaped by multiple mechanisms operating across hormonal, sleep-wake, behavioral and environmental factors.

This study is ancillary to an ongoing clinical trial and has limitations. Our sample size (n=5) limits generalizability to other brain regions and brain disorders. We cannot fully separate stimulation effects from natural rhythm dynamics, though we leveraged limited opportunities where stimulation was disabled in double-blinded settings for rigorous evaluation of the main findings. Additionally, self-reported symptom severity measures (ecological momentary assessment) have known limitations in mood disorders such as recall bias, state-dependent memory, and mood-congruent reporting^30,31^, although convergence of our results with clinician-guided MADRS allays these concerns. Future studies in larger TRD and non-TRD cohorts will be needed to determine the prevalence and clinical reach of multidien organization in mood disorders.

Our work reveals a promising new target for closed-loop neuromodulation based on the phase of endogenous, multidien rhythms that are pervasive in ongoing brain activity. Unlike existing paradigms in which stimulation reacts to instantaneous biomarkers, a phase-targeted paradigm would deliver therapy when the participant traverses a state most pliable to alleviating symptom onset, such as during the rising phase before peak depression. This protocol would align with chronotherapeutic principles established for light and sleep therapies^32^ for affective disorders, but never operationalised at the multidien scale with implanted devices. Multidien rhythms thus provide a forecastable, manipulatable temporal scaffold for mood, and a foundational principle on which precision psychiatry can be built.

## Methods

### Study participants

We recruited five adults (two male, three female) with treatment-resistant depression who had been implanted with the Responsive Neurostimulation System (NeuroPace, Inc), each with intracranial depth leads sampling two of the following regions: orbitofrontal cortex, amygdala, subgenual cingulate, hypothalamus, thalamus, hippocampus, nucleus accumbens, and anterior limb of the internal capsule, as part of the PRESIDIO trial (NCT04004169) (Extended Data Tables 1-2). Participants provided written informed consent for retrospective use of RNS System data. Experimental procedures were approved by the University of California, San Francisco (UCSF) Institutional Review Board (no. 17-23724).

### Data acquisition

#### Behavioral assessments

Participants were instructed to self-report mood on a Visual Analog Scale of Depression (VAS-D; 0-100 continuous scale) at least twice a day while swiping a magnet over their device to trigger storage of an electrocorticogram, thus providing a dense ambulatory readout of mood over time. The VAS-D was operationalized as an online survey with a horizontal slider on which participants could select any continuous point between 0 (not depressed at all) and 100 (worst depression). Scales were collected and managed using the secure and HIPAA-compliant web-based system Research Electronic Data Capture (REDCap). Additionally, the Montgomery-Asberg Depression Rating Scale (MADRS) was administered approximately every two weeks (mean:14.5, std.dev: 4.7, range: 9.5–19.8 days) to assess clinical depression severity.

#### RNS recordings

The RNS System provided four channels of intracranial EEG (iEEG), recorded as a bipolar derivation sampled at 250 Hz. Each recording was stored as a 240-s snapshot of the raw local field potential (LFP), comprising 160 s before and 80 s after the magnet swipe (2:1 ratio of before:after the swipe)^33^. Snapshots were stored when triggered by the participant swiping a magnet over the device (“magnet recordings”). Participants were instructed to swipe the magnet once, wait four minutes, submit an online VAS-D rating, and swipe the magnet a second time after the four minutes have elapsed or after completing the rating, whichever occurs later. Participants were asked to collect iEEG and VAS-D ratings twice a day, and any time they experience improvement or worsening of depression severity.

#### Determination of personalized stimulation and recording sites

Prior to chronic device implantation, each participant underwent a multi-day in-patient period of intracranial corticolimbic mapping to identify personalized stimulation and sensing targets, following a previously reported approach^15^. During this period, multi-day intracranial electrophysiology and focal electrical stimulation were used to identify a participant-specific neural biomarker of high-symptom state (defined as a neural frequency band in an implanted location showing highest correlation with VAS-D) and a stimulation location at which focal stimulation improved symptoms. This personalized mapping yielded one best stimulation-sensing target pair per participant, which guided placement of the chronic RNS System leads (Extended Data Table 2) and the subsequent delivery of neurostimulation.

#### Configuration of detection settings

After RNS implantation, two detection settings were identified for each participant - fixed-schedule stimulation, and closed-loop stimulation. For fixed-schedule stimulation, stimulation was delivered with a fixed duty cycle for approximately 12-14 hours daily, to reach the desired number of daily stimulations. Fixed-schedule stimulation was achieved using the responsive stimulation device by employing the ‘short episode treatment strategy’. By using a specific combination of settings, the detector was programmed with a sensitivity such that it was triggered continuously; the ‘post-episode interval’ was then used to control the interval between stimulations. With this combination, we were able to achieve fixed duty cycle stimulation. Closed-loop stimulation was controlled by each participant’s personalized neural biomarker, a neural activity pattern associated with high symptoms. For this detection setting, the device is programmed to be eligible for stimulation starting at the same time each day and stimulation is delivered when the device detection is triggered. Several detection parameters are adjusted to refine detection sensitivity in order to achieve the desired number of stimulations during hours of wakefulness. Both detection settings deliver stimulation at the same current, frequency, and electrode contacts.

### Data processing

#### Construction of daily time series

We constructed a daily time series of mood and neural data by averaging VAS-D scores and neural recordings over each calendar day, to match the temporal resolution of the analysis to the multidien frequencies of interest (periods ≥ 2 days). Each iEEG snapshot was subjected to a stimulation-artifact detector pipeline^34^ to identify stimulation artifacts, and 5 seconds of data around each stimulation event was removed. The remaining artifact-free recording was averaged to a single mean voltage value per channel. These per-snapshot mean voltages were then averaged across all snapshots acquired on the same calendar day to yield one iEEG value per day per recording channel. Similarly, VAS-D scores acquired on the same calendar day were averaged to get one mood score per day.

#### Preprocessing

We preprocessed the neural data and mood data to address specific confounds of longitudinal ambulatory recordings such as implantation effect and participants’ recording compliance. First, we discarded the first three months of iEEG recordings (and the corresponding days of mood data) in each participant to account for destabilization of neural signals right after implantation (implantation effect)^35^. Second, we addressed missing days of data (days in which participants might have failed to record data) by computing length (in days) of consecutive missing data, and partitioning the timeseries into segments wherever ≥12 consecutive days of data were missing. Shorter gaps (<12 d) were retained within a segment. Since we analyzed multidien rhythms starting from a period of 2 days, and looked up to 34 days using a 6-cycle wavelet, the longest permissible missing period length was fixed at 2 x 6 = 12 days. Additionally, segments shorter than 204 days (34 x 6 cycles; i.e. six cycles of the longest analyzed multidien period of 34 d) were discarded to ensure every analyzed segment was long enough to resolve the slowest multidien rhythm. Third, missing data gaps (<12 days) were interpolated using Kriging interpolation (based on prior reported use for multidien rhythm analysis in epilepsy^6^) since wavelet transform requires continuous data without gaps.

#### Detrending and normalization

Each interpolated time series (four iEEG channels and one VAS-D series per participant) was subjected to linear detrending to remove slow non-oscillatory drifts by subtracting a first-degree polynomial fit. Each detrended series was then z-scored to allow comparison across channels and participants. The VAS-D series was subjected to the same pipeline (daily averaging, segmentation, minimum-duration filtering, Kriging interpolation, linear detrending, and z-scoring).

#### Extraction of multidien rhythms

We performed wavelet-based time-frequency analysis on both the preprocessed iEEG time series and the VAS-D time series to extract time-varying multidien iEEG rhythms and mood cycles. Following the multidien wavelet framework established in epilepsy^6,7^, we used a broadband six-cycle complex Morlet wavelet transform, which provides an approximately even trade-off between time and frequency resolution and is the standard choice for resolving quasi-periodic biological rhythms. We constructed a family of wavelet scales spaced logarithmically^36^, with the smallest scale defined as 2 days, 20 scales per octave, and the largest scale corresponding to a period of 34 days. This yielded a family of 82 wavelets with periods spanning 2-34 days, densely tiling the multidien band. We convolved each wavelet with iEEG and VAS-D time series separately to extract complex-valued wavelet coefficients reflecting the amplitude and phase information of multidien rhythms and cycles for each multidien scale and time point. Multidien periodograms were computed as time-averaged wavelet power at each scale. The dominant period was the periodogram maximum, with a 95% CI from bootstrap resampling of days (2,000 resamples). Periodograms were normalised to unit sum and compared across participants by mean pairwise Pearson correlation, tested against a circular-shift null.

### Broadband multidien rhythm and clinical depression phase preference

#### Broadband rhythm approach

Summarizing a person’s momentary position in the cycle using any one narrowband multidien scale is limited by the fact that multidien rhythms are non-stationary quasi-rhythms that can fluctuate with multiple periods, change period length and undergo phase shifts^16,18^. Additionally, we observed highly individualized multidien rhythms and cycles in our participants. Therefore we chose to use a broadband approach over a narrowband approach when quantifying both multidien iEEG rhythms and mood cycles. We collapsed the full multidien band (2-34 days) into one broadband rhythm using an established broadband wavelet decomposition method^16,18^. This broadband rhythm’s amplitude and phase was then used for further analyses. Broadband amplitude envelopes were computed per channel as the modulus of the scale-averaged wavelet coefficient. Drift was quantified as the Pearson correlation between envelope and recording day.

#### Phase preference of depression severity

To test if momentary position in the mood cycle carries clinical information, we asked how VAS-D and MADRS score magnitude changed as a function of the broadband VAS-D phase. Each VAS-D and MADRS observation was assigned the broadband VAS-D phase of its day, and the scores were plotted as a function of the VAS-D phase in polar histograms with 40-degree phase bins (9 bins with 50% overlap spanning –π to +π). Phase preference was quantified as the score-weighted mean resultant vector, in which each day’s phase was weighted by its VAS-D or MADRS score after subtracting the participant’s minimum score to make weights non-negative, and whose length (R) gives strength of phase concentration and whose angle *θ* gives the preferred phase. Significance was assessed by permuting the scores (1000 permutations) and recomputing R; p = fraction with R ≥ observed R. At the cohort level, scores were permuted within each participant and the resulting R averaged across participants on each permutation, giving a null distribution for the mean R; p = fraction of this null ≥ the observed mean R.

### iEEG-mood multidien rhythm relationship

#### Coherence

Synchronization between multidien iEEG rhythms and mood cycles was measured by computing the magnitude coherence^37^, which measures the consistency of their phase relationship at each scale. For complex wavelet coefficients *W_N_* (neural) and *W_V_* (VAS-D), coherence at scale *s* is

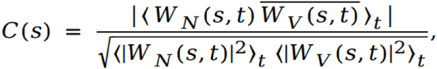

where the overline denotes complex conjugation and ⟨⋅⟩*t* denotes averaging over time. *C*(*s*) ranges from 0 (no consistent phase relationship) to 1 (perfect synchronisation). Coherence was computed per channel and averaged within each brain region. Significance was assessed against a null of shuffled VAS-D series, with the 95th percentile defining the threshold at each period. Peak coherence period was the maximum of each region’s coherence spectrum; regions were compared by Kruskal–Wallis test. For cohort-level analysis, coherence was averaged across all four iEEG channels per participant.

#### Time-varying correlation

Correlation between VAS-D and iEEG over time was calculated using a rolling window of 14 days (matched to the biweekly clinical depression assessment cadence) advanced in 1-day steps.

#### Phase difference and lead-lag

To quantify temporal ordering and lead-lag relationships between iEEG rhythms and mood cycles, we computed the instantaneous phase difference between the broadband iEEG rhythm and mood cycle for each participant as

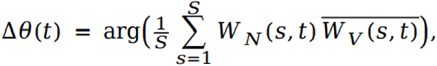

where a positive Δ*θ*(*t*) indicates that the neural rhythm is phase-advanced relative to the mood cycle (i.e. neural rhythm is leading the mood cycle). For each participant, Δ*θ*(*t*) was pooled across the four iEEG channels and over time, and we summarised the distribution by its circular mean (µ) and resultant vector length (R) defined as

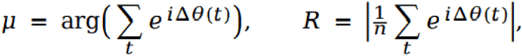

where µ quantifies the phase difference between the two rhythms, and R quantifies phase consistency. A 95% confidence interval on µ was obtained by a circular bootstrap over days (3,000 resamples); a neural lead over mood cycle was considered statistically significant when this interval excluded zero in the positive direction.

### Forecasting models

All forecasting models were trained per participant and evaluated on held-out data separated from training data by a purging gap to prevent train-test information leakage. For a target series *y* and forecast horizon *H* (days), the model was trained to predict *y*(*t* + *H*) from the wavelet features at day t (a single feature vector per day across all scales), with performance reported as the Pearson correlation *r* between observed and predicted values on the test set (scalar targets) or as the mean resultant vector *R* of prediction errors (phase targets). Significance was assessed against a permutation null obtained by shuffling the target.

#### Train-test split with purging

We took the first 70% of the data as the training set, and implemented a purging gap between the training and test to prevent train-test data leakage arising from the nature of wavelet convolution. Wavelet convolution using a centered wavelet of scale s at day t computes the wavelet coefficient at day t using information from t-x/2 to t+x/2 days (where x is the width of the wavelet). This property leads to data leakage at the train/test boundary, where the longest-scale wavelet coefficients on the training side draw on data from the test side, which would leak information across the split. To prevent this, we separated the training and test sets by a purging gap defined as a buffer of *P* days immediately following the training window, excluded from both the training and test sets. The purging gap width *P* was set to the wavelet e-folding time of the longest scale, evaluated as the lag at which the magnitude of the Morlet wavelet’s envelope decays to 1/*e* of its peak^36^, equal to √2 *s* for scale *s*. Evaluated at the largest scale (34 days), this gave *P* ≈ 47 days. Thus, for a time series of *N* days and a training fraction *f*, the model was trained on the first *fN* days; the following *P* days were discarded as a purging gap, belonging to neither set; and the model was tested on the remaining *N*−*fN*−*P* days.

### Forecasting mood from mood rhythm

To test which property of the mood rhythm best predicts future mood, we compared three feature sets as predictors of VAS-D score. From the VAS-D wavelet decomposition, we formed at each day *t*:

(i) phase features: the sine and cosine of the phase angle of the mood cycle at every scale S as

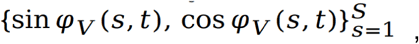

in order to project the circular phase at each scale into Cartesian coordinates so that it can enter a linear model;

(ii) amplitude features: the amplitude at every scale as

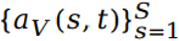

and

(iii) score: the raw daily VAS-D value, *V*(*t*). Each feature set was used to fit a separate Ridge regression model predicting VAS-D at horizon *H*. The phase model was defined as

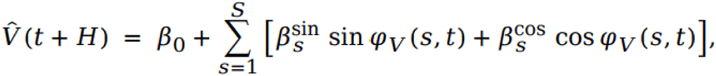

the amplitude model as

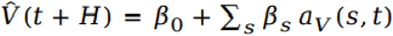

and the score model as

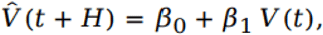

respectively, where *β*_0_ is the model intercept and the *β* are the weights the model learns for each feature and encodes the relative contribution of the feature value at each multidien scale towards predicting future mood.

Coefficients were estimated by minimizing the penalized loss

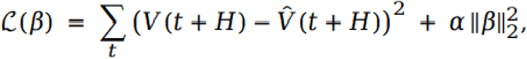

where the Ridge penalty *α* was selected by 5-fold cross-validation within the training set over a logarithmic grid (*α* ∈ [10^−2^, 10^6^]). To characterize the model’s forecasting performance (Pearson’s *r* between predicted and observed VAS-D scores) over different horizons, we repeated the analysis for horizons *H* = 1, … , 30 days. To evaluate the model’s performance as a function of training data length, we repeated the analysis by varying the training fraction *f* from 0.50 to 0.75 and measured performance (Pearson’s *r*) as a function of training fraction.

### Forecasting mood cycle phase from neural phase

Ridge-regression models were trained for each participant using the multidien phase of intracranial neural activity (all four channels combined) to predict the broadband phase of the mood (VAS-D) multidien cycle one day ahead. Features at day t, X(t), consisted of the sine and cosine of the neural phase Ψ at every scale (s) and channel (c),

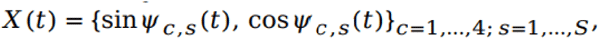

calculated for all four iEEG channels. Since the target (mood phase) is circular, we predicted its sine and cosine with two Ridge regression models^38^ defined as

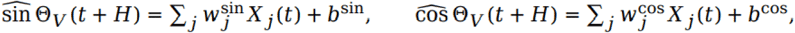

and the predicted phase was recovered as

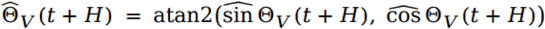

after normalizing the predicted sine and cosine to unit length. Ridge penalties were selected by cross-validation within the training set over a logarithmic grid: *α* ∈ [10⁻², 10³] (25 values, leave-one-out generalised cross-validation). Phase-prediction accuracy was quantified as the mean resultant vector R of the phase differences between predicted and observed phase

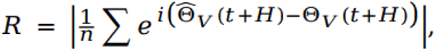

where R = 1 denotes perfect phase tracking, while R = 0 denotes chance, with significance assessed against a null distribution of R obtained by randomly permuting the observed phases.

Models were evaluated across horizons *H* = 1, … , 30 days. Per-participant significance was assessed by permuting the observed VAS-D phases against the forecasted phases and recomputing R (100 permutations); p = fraction of surrogate R ≥ observed. At the cohort level, the group statistic reported as the mean across-participant R, tested against a null formed by averaging each participant’s permutation null across participants (p = fraction of this null ≥ the observed mean R).

As a control, we tested whether forecasting performance depended specifically on the wavelet-derived multidien phase rather than on the neural signal itself. We refit an identical model in which the phase features were replaced by the raw iEEG voltage - i.e. we used the iEEG voltage directly without wavelet decomposition or phase extraction. The feature vector at day t was defined as

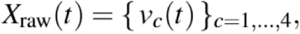

where v_c_(t) is the raw iEEG voltage calculated for all four iEEG channels (c=4).

As additional baselines, we trained two further models on the same target and pipeline. A VAS-D score model was trained using the participant’s raw daily VAS-D score to test whether the mood score alone could predict future mood-cycle phase without any neural information. A combined VAS-D + iEEG voltage model was trained, whose features concatenated the daily VAS-D score with the same-day raw iEEG voltage from all four channels. This tested whether behavioural and raw neural signals together (without wavelet-phase extraction) could match the multidien phase model. Every other component of the pipeline was identical to the phase model: the broadband multidien mood cycle phase was the prediction target; its sine and cosine were predicted with two cross-validated Ridge models and recovered after unit-normalization; and the mean resultant vector R, with its per-participant and cohort-level permutation nulls, was computed across horizons *H* = 1, … , 30 days. Differences in R between these three baseline models and the multidien phase model would therefore isolate the predictive contribution of the multidien-phase representation.

### Effect of stimulation on multidien rhythms

We analyzed how multidien rhythms differed between stimulation on and stimulation off periods (each of multiple weeks’ duration). In contrast to prior studies that examine the acute effects of stimulation, we analyzed the cumulative effects of stimulation events over multiple days.

#### Stimulation conditions

Only three of the five participants completed a randomised, blinded cross-over study with three arms: sham, in which the device recorded but delivered no stimulation; closed-loop (CL) stimulation arm, in which the device continuously monitored a pre-specified neural biomarker and delivered stimulation only when that biomarker crossed a set threshold (responsive, biomarker-triggered stimulation); and intermittent (INT) stimulation arm, in which stimulation was delivered on a fixed schedule, independent of any biomarker. Arm durations were 42 days per arm for S2, 83–91 days for S4, and 76-84 days for S5. We referred to sham days as stimulation-off and CL and INT days as stimulation-on.

#### Wavelet convolution

For each arm, we re-computed a wavelet decomposition of the daily series restricted to that arm. The largest scale within each arm was set to *s*_max_ = (arm length in days)/ *ω*_0_ (where *ω*_0_ is 6 since we used a 6-cycle wavelet convolution) so that each arm’s wavelet spectrum was calculated only for the scales that could be resolved within that arm. Critically, each arm was transformed independently and analyzed only over scales resolvable within that arm, so that the longer multidien scales did not draw on data spanning across arms.

#### Rhythm amplitude

We quantified the strength of the multidien neural rhythm expressed within each arm by computing the time-averaged power ⟨|*W*(*s*, *t*)|^2^⟩*_t_* across multidien scales for each arm, and comparing the multidien power between the arms using a paired t-test across participants.

#### Phase occupancy

To test whether stimulation affected amount of time spent in each phase of the neural rhythm, we classified each day by its position on the broadband cycle into one of four equal 90^∘^ quadrants (rising, peak, falling and trough) and computing the percentage of days falling in each quadrant for each arm. Quadrants were then pooled into two groupings: the peak-depression phase and the transition phases (rising and falling). For each participant we computed the stimulation-induced change in phase occupancy as the difference from sham for each active arm (CL - sham and INT - sham); these differences were pooled across the three participants and two active conditions (*n* = 6) and the change in extreme-phase occupancy was compared with the change in transition-phase occupancy using a paired *t*-test.

### Statistics

Significance was assessed for primary analyses by comparing against permutation or circular-bootstrap null distributions. Permutation p-values were computed as p = (surrogates ≥ observed + 1)/(permutations + 1). For cohort-level tests, the observed statistic was averaged across participants and compared against a null formed by averaging each participant’s surrogate distribution. Circular means and resultant vector lengths were used for phase quantities, and confidence intervals were computed from circular bootstrap (2,000–3,000 resamples).

Group-level comparisons across participants were tested using non-parametric two-sided tests: Kruskal–Wallis for peak coherence period across regions, Friedman for accuracy across channel-subset sizes, Wilcoxon signed-rank for pairwise model comparisons, and paired t-tests for stimulation-arm contrasts.

Analyses used Python 3.11.5 with NumPy, SciPy, pandas, scikit-learn, statsmodels, pingouin, pycircstat, pycwt, pyeisen, PyKrige, Astropy, Matplotlib and seaborn. Random seeds were fixed, to make permutation and bootstrap results reproducible.

## Acknowledgements

This work was supported by the Ray and Dagmar Dolby Family Fund through the Department of Psychiatry at UCSF and by the National Institute Of Neurological Disorders And Stroke of the National Institutes of Health under Award Number UH3NS123310. The content is solely the responsibility of the authors and does not necessarily represent the official views of the National Institutes of Health. We thank Krishnakant Saboo for valuable discussions during development of this study.

## Author information

### Contributions

N.P.S., A.N.K., A.D.K, and E.F.C. initiated the work and supervised the study. J.F., K.K.S., D.A., A.T., N.B., A.A., E.H., A.L.B., and L.P.S. collected the data. N.P.S., A.N.K., K.K.S., and J.M.F. analyzed the data and N.P.S. and A.N.K. drafted the manuscript. A.N.K., E.F.C., and A.D.K. finalized the manuscript. All authors approved the work and take responsibility for its integrity.

## Ethics declarations

### Data availability

Raw RNS System data were obtained through a Data Use Agreement between UCSF and NeuroPace and therefore cannot be publicly shared.

## References

1. Barrett, L. F., Mesquita, B., Ochsner, K. N. & Gross, J. J. The Experience of Emotion. Annu. Rev. Psychol. 58, 373–403 (2007).

2. Frank, E. Conceptualization and Rationale for Consensus Definitions of Terms in Major Depressive Disorder: Remission, Recovery, Relapse, and Recurrence. https://jamanetwork.com/journals/jamapsychiatry/fullarticle/495503 (1991).

3. Solomon, D. A. et al. Multiple recurrences of major depressive disorder. Am. J. Psychiatry 157, 229–233 (2000).

4. Silva, R. I. D. et al. Multiday rhythms modulate human heart rate: an observational study in healthy adults. 2026.03.01.708870 Preprint at 10.64898/2026.03.01.708870 (2026).

5. Ceolini, E. & Ghosh, A. Common multi-day rhythms in smartphone behavior. Npj Digit. Med. 6, 49 (2023).

6. Baud, M. O. et al. Multi-day rhythms modulate seizure risk in epilepsy. Nat. Commun. 9, 88 (2018).

7. Leguia, M. G. et al. Seizure Cycles in Focal Epilepsy. JAMA Neurol. 78, 454–463 (2021).

8. Karoly, P. J. et al. Cycles in epilepsy. Nat. Rev. Neurol. 17, 267–284 (2021).

9. Gregg, N. M. et al. Seizure occurrence is linked to multiday cycles in diverse physiological signals. Epilepsia 64, 1627–1639 (2023).

10. Benedetti, F., Barbini, B., Colombo, C., Campori, E. & Smeraldi, E. Infradian mood fluctuations during a Major Depressive episode. J. Affect. Disord. 41, 81–87 (1996).

11. Wehr, T. A. Bipolar mood cycles and lunar tidal cycles. Mol. Psychiatry 23, 923–931 (2018).

12. Alagapan, S. et al. Cingulate dynamics track depression recovery with deep brain stimulation. Nature 622, 130–138 (2023).

13. Fitoz, E. C. et al. Common Electrophysiology Biomarkers Collected at Home Robustly Track Depression Recovery With Deep Brain Stimulation. MedRxiv Prepr. Serv. Health Sci. 2026.04.13.26350107 (2026) doi:10.64898/2026.04.13.26350107.

14. Scangos, K. W., Makhoul, G. S., Sugrue, L. P., Chang, E. F. & Krystal, A. D. State-dependent responses to intracranial brain stimulation in a patient with depression. Nat. Med. 27, 229–231 (2021).

15. Scangos, K. W. et al. Closed-loop neuromodulation in an individual with treatment-resistant depression. Nat. Med. 27, 1696–1700 (2021).

16. Leguia, M. G., Rao, V. R., Kleen, J. K. & Baud, M. O. Measuring synchrony in bio-medical timeseries. Chaos Interdiscip. J. Nonlinear Sci. 31, 013138 (2021).

17. Proix, T. et al. Forecasting seizure risk in adults with focal epilepsy: a development and validation study. Lancet Neurol. 20, 127–135 (2021).

18. Khambhati, A. N., Chang, E. F., Baud, M. O. & Rao, V. R. Hippocampal network activity forecasts epileptic seizures. Nat. Med. 30, 2787–2790 (2024).

19. Kirkby, L. A. et al. An Amygdala-Hippocampus Subnetwork that Encodes Variation in Human Mood. Cell 175, 1688–1700.e14 (2018).

20. Sani, O. G. et al. Mood variations decoded from multi-site intracranial human brain activity. Nat. Biotechnol. 36, 954–961 (2018).

21. Provenza, N. R. et al. Disruption of neural periodicity predicts clinical response after deep brain stimulation for obsessive-compulsive disorder. Nat. Med. 30, 3004–3014 (2024).

22. Mayberg, H. S. et al. Deep brain stimulation for treatment-resistant depression. Neuron 45, 651–660 (2005).

23. Holtzheimer, P. E. et al. Subcallosal cingulate deep brain stimulation for treatment-resistant depression: a multisite, randomised, sham-controlled trial. Lancet Psychiatry 4, 839–849 (2017).

24. Chiang, S. et al. Evidence of state-dependence in the effectiveness of responsive neurostimulation for seizure modulation. Brain Stimulat. 14, 366–375 (2021).

25. Wong, S. N., Halaki, M. & Chow, C. M. The periodicity of sleep duration – an infradian rhythm in spontaneous living. Nat. Sci. Sleep 5, 1–6 (2013).

26. Reinberg, A. E., Dejardin, L., Smolensky, M. H. & Touitou, Y. Seven-day human biological rhythms: An expedition in search of their origin, synchronization, functional advantage, adaptive value and clinical relevance. Chronobiol. Int. 34, 162–191 (2017).

27. Pariante, C. M. & Lightman, S. L. The HPA axis in major depression: classical theories and new developments. Trends Neurosci. 31, 464–468 (2008).

28. Nicolaides, N. C., Charmandari, E., Chrousos, G. P. & Kino, T. Circadian endocrine rhythms: the hypothalamic–pituitary–adrenal axis and its actions. Ann. N. Y. Acad. Sci. 1318, 71–80 (2014).

29. Bonsall, M. B., Geddes, J. R., Goodwin, G. M. & Holmes, E. A. Bipolar disorder dynamics: affective instabilities, relaxation oscillations and noise. J. R. Soc. Interface 12, 20150670 (2015).

30. Colombo, D. et al. Current State and Future Directions of Technology-Based Ecological Momentary Assessment and Intervention for Major Depressive Disorder: A Systematic Review. J. Clin. Med. 8, 465 (2019).

31. Wenze, S. J. & Miller, I. W. Use of ecological momentary assessment in mood disorders research. Clin. Psychol. Rev. 30, 794–804 (2010).

32. Wirz-Justice, A. et al. Chronotherapeutics (light and wake therapy) in affective disorders. Psychol. Med. 35, 939–944 (2005).

33. Sellers, K. K. et al. Closed-Loop Neurostimulation for Biomarker-Driven, Personalized Treatment of Major Depressive Disorder. J. Vis. Exp. JoVE 10.3791/65177 (2023) doi:10.3791/65177.

34. Khambhati, A. N., Shafi, A., Rao, V. R. & Chang, E. F. Long-term brain network reorganization predicts responsive neurostimulation outcomes for focal epilepsy. Sci. Transl. Med. 13, eabf6588 (2021).

35. Sun, F. T., Arcot Desai, S., Tcheng, T. K. & Morrell, M. J. Changes in the electrocorticogram after implantation of intracranial electrodes in humans: The implant effect. Clin. Neurophysiol. Off. J. Int. Fed. Clin. Neurophysiol. 129, 676–686 (2018).

36. Torrence, C. & Compo, G. P. A Practical Guide to Wavelet Analysis. Bull. Am. Meteorol. Soc. 79, 61–78 (1998).

37. Bruña, R., Maestú, F. & Pereda, E. Phase locking value revisited: teaching new tricks to an old dog. J. Neural Eng. 15, 056011 (2018).

38. Sarma, Y. and Jammalamadaka, S. Circular Regression. Statistical Science and Data Analysis, 109-128. Proceeding of the Thrid Pacific Area Statistical Conference. VSP: Utrecht, Netherlands. (1993).

